# Study protocol and statistical analysis plan for a randomized controlled trial evaluating the safety and feasibility of the recombinant human platelet-derived growth factor B (rhPDGF-BB)-enhanced collagen plug for complex perianal fistula healing

**DOI:** 10.64898/2026.06.22.26356267

**Authors:** Marissa C Kuo, Samuel A Younan, Melissa D Lempicki, Alex T Hawkins, Jacob Smith, Jana K Shirey-Rice, Jill M Pulley, Samuel E Lynch, Thomas E Ueland, Aimal Khan, Cyndi R Clark, Bryan S Blette

## Abstract

**Background:** A drug-repurposing-specific phenome-wide association study (PheWAS) demonstrated that patients with a single nucleotide variant that decreases expression of platelet-derived growth factor receptor beta (PDGFRβ) have a higher prevalence of fistulas, suggesting that PDGFRβ signaling is important for tissue repair. Recombinant human platelet derived growth factor B (rhPDGF) is an FDA-approved protein-based therapeutic that signals through PDGFRβ to heal and regenerate cutaneous skin wounds, periodontal tissue, and orthopedic bone with a strong safety profile. We hypothesize that rhPDGF will benefit other conditions identified by PheWAS with a similar physiological mechanism as the existing indications, such as complex perianal fistulas that are ineligible for a fistulotomy.

**Methods and analysis:** This prospective, blinded, single-site study aims to enroll 12 participants, randomized at a ratio of 2:1, comparing implantation of rhPDGF-enhanced collagen to routine care procedures, and stratified by fistula etiology, idiopathic versus Crohn’s disease (CD)-related. The primary outcome of this study will evaluate the technical performance of the rhPDGF-enhanced collagen implant for treatment of complex perianal fistulas as measured by the proportion of participants with successful implantation of the intervention without any intervention-related serious adverse events. The secondary outcomes will assess the preliminary safety and efficacy of the intervention based on all intervention-related adverse events, total fistulas healed, rate of fistula recurrence, and change in patient-reported symptoms. Complex perianal fistulas, idiopathic or CD-related, remain a major clinical challenge in need of new multimodal treatments aimed at tissue repair and regeneration. Pharmaceutical rhPDGF stimulation of PDGFRβ signaling promotes healing of skin, bone, and soft tissue. PheWAS revealed fistulas as a novel indication for repurposing rhPDGF. This protocol aims to evaluate the technical performance, preliminary safety and efficacy, and feasibility of rhPDGF-enhanced collagen for healing and remission of complex perianal fistulas.

**Ethics and dissemination:** This trial was approved by the Vanderbilt University Medical Center institutional review board (IRB#240585). Results will be submitted for publication in a peer-reviewed journal.

**Clinical trial registration:** This trial is registered at ClinicalTrials.gov (NCT06632418).

**STRENGTHS AND LIMITATIONS OF THIS STUDY:**

- In this trial we utilized a novel drug repurposing approach, PheWAS, to identify fistulas as a novel pathophysiological phenotype which may benefit from a pharmacological intervention targeting the PDGF signaling pathway.
- A crossover design was employed to improve recruitment and retention, allowing patients within the control group the opportunity to receive the intervention if their fistula fails to heal.
- Our secondary outcomes will include stratified subgroup analyses for idiopathic and CD-related fistulas allowing us to determine if PDGF may benefit one fistula etiology over the other.
- Due to the complexity of perianal fistula etiology, stringent exclusion criteria were required to establish a study population who might benefit the most and the small sample size restricts the ability to perform subgroup analyses and limits statistical power for efficacy outcomes.
- The control procedures differ based on disease etiology (idiopathic vs CD-related) yet all patients undergoing treatment receive the same procedure, as such, the interpretation of the secondary outcomes may be difficult due to the lack of a true placebo.

## INTRODUCTION

Repurposing drugs with existing safety data has many advantages in clinical trials but involves the identification of a novel and precise indication where the drug can address an unmet medical need effectively. Vanderbilt University Medical Center’s (VUMC) academic drug repurposing program leverages human genetic variation to inform drug repurposing and bypass the limitations of *in vitro* and preclinical models. Phenome-wide association studies (PheWAS) using deidentified patient DNA samples, single nucleotide variant (SNV) genotyping data, and electronic health records can identify and validate associations between SNVs that cause specific protein changes that correlate with a drug’s effects and novel disease phenotypes that are potentially treatable by the drug (1–3). A PheWAS analysis performed by our team linked a SNV in the platelet-derived growth receptor beta gene (*PDGFRB),* that decreases PDGFRβ protein expression, with *increased* prevalence of chronic wounds, tissue defects, and higher rates of graft, reconstructive, and transplant procedures in patients carrying at least one copy of the variant allele. This suggests that the PDGFRβ signaling pathway is critical during wound healing and tissue regeneration following tissue injury or damage and that stimulation of this pathway may accelerate healing and promote repair (4).

PDGFRβ is one of two isoforms of the tyrosine kinase receptors that bind the PDGF family of growth factors found naturally in the body. PDGF-BB homodimers bind to PDGFRβ and promote chemotaxis, mitogenesis, and survival of many different cell types and extracellular matrix deposition during embryonic development and tissue repair following injury. Recombinant human PDGF-BB (rhPDGF) is a pharmaceutical recombinant protein therapy that mimics the activity of the endogenous growth factor that is FDA-approved for the treatment of diabetic foot ulcers (DFU), periodontal bone and soft tissue reconstruction, and orthopedic bone regeneration, and has over 150 published clinical studies demonstrating its safety. Our PheWAS analysis showed an increased prevalence of DFUs, periodontitis, and orthopedic bone disorders associated with the PDGFRβ SNV. The presence of these existing indications in the dataset validated the biological consequences of the SNV and supported the selection of a novel phenotype from the PheWAS. Furthermore, the analysis revealed an increased prevalence of fistula occurrence associated with the PDGFRβ SNV, revealing a novel indication for repurposing rhPDGF.

Perianal fistula, an abnormal connection between the rectum and perianal epithelium, is a common cause of anorectal burden with an overall incidence of 9 cases per 100,000 (5). Anal fistulas account for up to 5% of proctological consultations and are often responsible for substantial detriment to patient quality of life in terms of pain, fecal incontinence, and psychologic distress (6). The generally accepted pathophysiology of idiopathic, or cryptoglandular, fistulas, accounting for 90% of all cases, includes obstruction of an anal gland followed by chronic infection and epithelization of the abscess drainage tract (7,8). However, mechanistic clarity regarding proposed pro-inflammatory and infectious factors remains elusive (9–12). The second most common cause of perianal fistulas is due to Crohn’s disease (CD), a chronic inflammatory disease of the gastrointestinal tract with an estimated annual incidence of 3 to 20 cases per 100,000 (13). Approximately 20% of patients with CD will develop a perianal fistula within 10 years of diagnosis (14) as a severe complication of chronic intestinal inflammation, extracellular matrix remodeling, and pathogen-associated molecular patterns resulting in epithelial-to-mesenchymal transition and tissue remodeling mechanisms that pave the fistula tract (15).

Idiopathic perianal fistulas are primarily treated with surgical interventions after seton placement which allows the abscess to drain and clears the infection. CD-related perianal fistulas rely on a combination of pharmacological therapies to reduce chronic inflammation and control infection in addition to seton placement, and definitive surgical procedures are only considered once inflammation is controlled. The optimal treatment strategy and prognosis depend on the fistula classification as simple or complex as defined by the anatomy, single tract or branching, and the proportion of the sphincter muscle that is involved (16). For patients with preoperative fecal continence and simple fistulas with minimal sphincteric involvement, fistulotomy, a procedure wherein the physician cuts open the length of the fistula, is often an effective treatment resulting in healing for over 90% of patients (17). Options for complex fistulas involving more of the sphincter muscle are broad and must be carefully selected to minimize the risk of long-term incontinence. Fistulotomy, fistulectomy, ligation of the intersphincteric fistula tract (LIFT), endorectal advancement flap, modified Hanley procedure, anal fistula plug implantation, or natural healing are the standard treatment choices with variable success rates between 15.8-72.7% for complete healing, likelihood of recurrence, and risk of long-term bowel incontinence (18,19).

Non-invasive, pharmacological, medical options are lacking for perianal fistulas (11,20). Recent investigations have focused on targeting the processes of collagen remodeling and epithelial to mesenchymal transformation to promote natural healing with mixed evidence regarding the effectiveness of implantable bioprosthetic collagen plug, acellular crosslinked collagen matrix, and fibrin glue scaffolds (8,21). Although these minimally invasive therapies lack clear superiority over traditional surgical techniques, the dearth of less invasive options and need for sphincter-sparing modalities justifies the continued search for and optimization of therapies that target the underlying mechanistic drivers of perianal fistula formation, persistence, and recurrence.

Increased prevalence of a fistula diagnosis was found in the PDGFRβ PheWAS dataset. As such, we hypothesized that targeting the PDGFRβ signaling pathway by delivery of rhPDGF will promote repair and prevent recurrence of complex perianal fistulas. Systematic review of the literature revealed that blood-derived products, concentrated with PDGF-BB, improved healing rates of various fistula types in preclinical and clinical studies (22–28), further supporting that targeting this biological mechanism may benefit complex perianal fistula treatment outcomes, addressing a major unmet need in the clinic. We propose that rhPDGF will enhance healing of perianal fistulas by stimulating chemotaxis and mitogenesis of the cell types that deposit connective tissue, form new blood vessels, and regrow damaged tissue. This clinical trial aims to evaluate rhPDGF as an adjunct therapy to the implantation of a collagen matrix for treatment of complex perianal fistulas not amenable to fistulotomy.

## METHODS

### Trial design

This study will be carried out in accordance with International Conference on Harmonisation Good Clinical Practice (ICH GCP) and the United States (US) Code of Federal Regulations (CFR) applicable to clinical studies (45 CFR Part 46, 21 CFR Part 50, 21 CFR Part 56, 21 CFR Part 312, and/or 21 CFR Part 812). This Phase II clinical trial will evaluate the performance, preliminary safety and efficacy, and feasibility of rhPDGF-enhanced collagen on healing complex perianal fistulas that are not eligible for a fistulotomy in patients 21 years of age and older. This prospective, blinded, single-site study is expected to enroll and randomize 12 participants at a 2:1 intervention versus control allocation ratio stratified by fistula etiology, idiopathic versus CD-related. The two arms will compare implantation of the rhPDGF-enhanced collagen to routine care procedures, which include COOK Biotech’s Biodesign^®^ Anal Fistula Plug implant for idiopathic fistulas or draining seton removal and natural healing for CD-related fistulas. Following the baseline procedure, patients will be evaluated at Months 1, 3, and 6 to assess healing of the external fistula opening and change in pain, incontinence, and other perianal fistula-related symptoms. Patients randomized to the control group who fail to heal at Month 3 will have the option to crossover to the intervention group and receive the rhPDGF-enhanced collagen intervention. The Standard Protocol Items: Recommendations for Interventional Trials (SPIRIT) schedule of enrollment, intervention, and assessments is included as **Figure 1**. A graphic study timeline of events is provided in **Figure 2**. The SPIRIT checklist is included in **S1 Appendix**. A full accounting of evaluations and unabridged protocol approved by the IRB is available in **S2 Appendix** (December 13, 2024; version 6.1) and important protocol modifications will be available from the corresponding author and by viewing the ClinicalTrials.gov (NCT06632418) study entry. A sample consent form is included in **S3 Appendix**.

**Figure 1.**
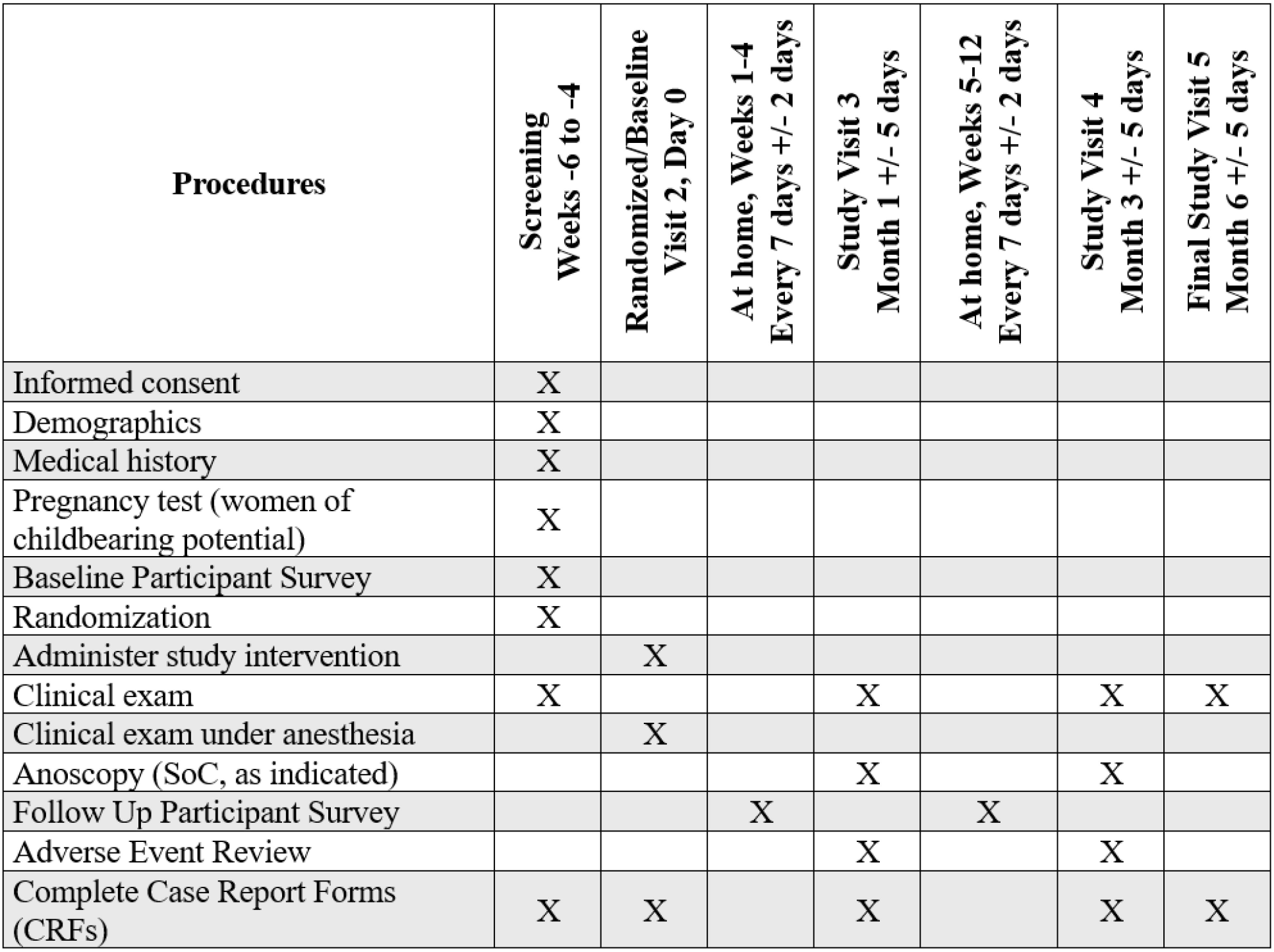
SPIRIT schedule of enrollment, intervention, and assessments

**Figure 2.**
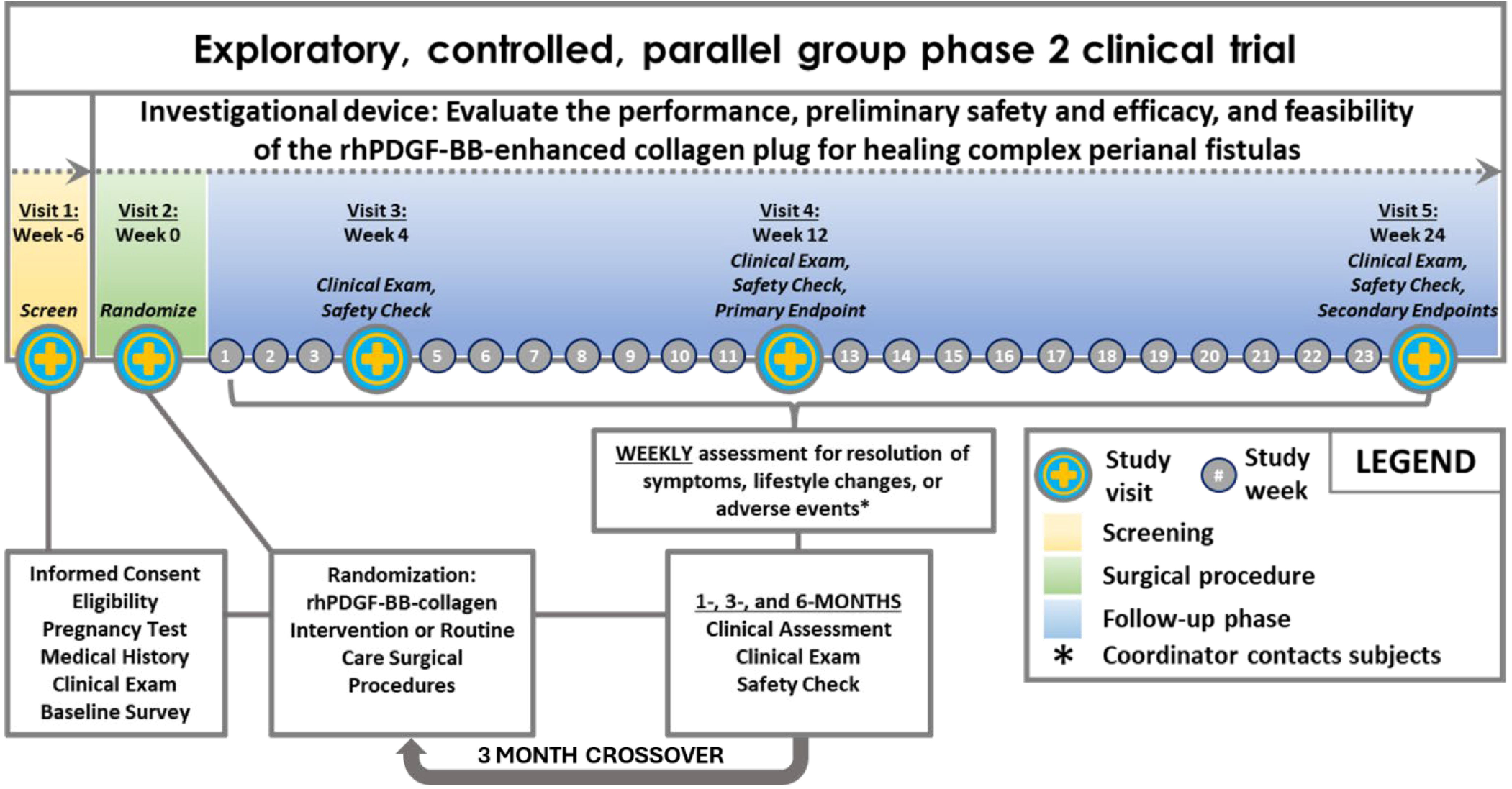
Schematic of study design

### Setting

The trial will be conducted at Vanderbilt University Medical Center (VUMC) in Nashville, TN in the Department of Surgery, Division of General Surgery, section of Colon and Rectal Surgery.

### Recruitment and Consent

This study aims to recruit 12 participants, that are adults >21 years of age regardless of gender, race, or ethnicity. There is no information currently available regarding differential agent effects in participants defined by race or ethnicity. No vulnerable populations are expected to be enrolled in this study. The target number of patients to be screened for enrollment is 60 with the expectation of enrolling and randomizing ∼25% of those screened and accounting for discontinuation, withdrawal, and loss to follow-up. This will be a single-site study at VUMC with an anticipated accrual rate of 1-2 participants per month.

Existing patients, new patients, and provider-referred patients with a diagnosis of a complex perianal fistula that is not amenable to a fistulotomy can be recruited for this trial. Patients that are interested in the trial will be scheduled for a screening visit where they will sign written informed consent and undergo the baseline examination for eligibility. Informed consent is initiated prior to the individual’s agreeing to participate in the study and continues throughout the individual’s study participation. Informed Consent Forms (ICF) will be VUMC Institutional Review Board (IRB)-approved. Only those participants who undergo the baseline procedure will be considered treated and count towards total participant population. Participants who signed consent but did not receive treatment will not be followed further.

### Participants

#### Inclusion criteria

1. Diagnosis of a single tract perianal fistula not amenable to fistulotomy as determined by the supervising surgeon
2. Fistula must currently have a draining seton in place
3. Aged >21 years old
4. Willing and able to provide informed consent for study participation and compliance with study protocol
5. Stated willingness to comply with all study procedures and availability for the duration of the study

#### Exclusion Criteria

1. Medical conditions that would, in the opinion of the investigator or treating provider, compromise the safety of the individual with study participation and/or the ability of the individual to follow study protocol
2. Genito-urinary fistulization, including rectovaginal (i.e., fistulas that transverse the vaginal canal)
3. Presence of an ileal anal pouch
4. Any major surgery of the gastrointestinal tract (including one or more segments of the colon or terminal ileum) within 3 months prior to randomization; presence of stoma is not exclusionary
5. Prior surgical procedure (i.e., Ligation of Intersphincteric Fistula Tract or Endorectal Advancement Flap) for the target fistula or a perianal procedure that resulted in a large soft tissue defect within 6 months prior to screening visit
6. One or more of the following fistula types or anatomic presentations: horseshoe fistulas, fistulas that do not have an opening inside the anal canal or low rectum, blind ending sinus tracts (no external opening), branching fistulas (a previously performed conversion of a branching fistula tract to a single tract is not exclusionary), >1 internal opening, moderate or severe proctitis, severe rectal mucosal fibrosis surrounding the internal opening preventing the securing of the fistula plug disk, any anatomical limitation to successfully securing the fistula plug disk.
7. Known allergic reactions to porcine collagen or yeast-derived products
8. Currently enrolled in a drug or device trial or within 30 days of last investigational drug or device administration at baseline visit where investigational treatment (drug or device) was placed in or near the fistula tract or may potentially interact with study treatment
9. Women who are known to be pregnant, breastfeeding, or planning to become pregnant, during the trial
10. Active infection at the application site
11. The presence of malignant neoplasms at the application site
12. Prior radiation therapy at the application site

#### Screen Failures

Screen failures are defined as participants who consent to participate in the clinical trial but are not subsequently randomized or the device is unable to be implanted. A minimal set of screen failure information is required to ensure transparent reporting of screen failure participants, to meet the Consolidated Standards of Reporting Trials (CONSORT) publishing requirements and to respond to queries from regulatory authorities. Minimal information includes demography, screen failure details, eligibility criteria, and any serious adverse event (SAE).

Individuals who consent and do not meet the criteria for participation in this trial (screen failure) because of the following criteria may be rescreened once the below criteria have been resolved.

- No draining seton in place
- Active infection
- Surgical procedures outlined in the exclusion criteria
- Participation in an investigational study as outlined in exclusion criteria
- Pregnant or breastfeeding

### Randomization and Blinding

Randomization will occur at the baseline visit at a ratio of 2:1 intervention versus control. Randomization will be stratified to achieve balance in treatment allocation across idiopathic and CD-related fistula types. Because the total sample size is 12, randomization will be blocked using blocks of size 3 (2:1 intervention to control) within each stratum (idiopathic versus CD-related). Based on historical clinical data we anticipate enrolling 9 Chron’s participants and 3 idiopathic participants. Blinding of the patients, surgeons other than the investigator performing the procedure, and study team members assessing outcomes to study arm allocation will be employed to reduce bias in conducting study activities and evaluations. Given that the control group will be receiving a routine care procedure, the investigator who will be performing all procedures will be unblinded to the treatment assignment for each participant. To maintain blinding while assessing outcomes, clinical exams during follow-up study visits will be performed by another surgeon not involved in the initial screening visit or the procedural encounter. In the case of a serious adverse event or medical emergency in which knowledge of the treatment is necessary for the participant’s well-being or medical care, the unblinded investigator will be alerted to ensure appropriate medical management.

### Intervention

rhPDGF is highly purified, aseptically processed, and sterile filled into transparent glass syringes. One syringe contains a 0.5 mL solution of 0.3 mg/mL rhPDGF in 20 mM sodium acetate buffer pH6+/-0.5. rhPDGF is stored at 2°C to 8°C and then allowed to reach room temperature prior to administration to the patient. Up to 4 mL of the rhPDGF (0.15-1.2 mg) solution will be used to rehydrate a collagen matrix manufactured from porcine collagen sourced from veterinary pigs. The native collagen molecules and fiber structures are maintained, and no additional materials are added. Antigens, lipids, and non-collagenous proteins are removed through extraction and purification steps rendering the material acellular and reducing the likelihood of acute and chronic biological reactions. The collagen membranes are available in three different sizes: 20×30 mm (model 500662), 30×40 mm (model 500663), and 40×50 mm (model 500664) and can be trimmed as needed. All components of the intervention are biocompatible and absorbable and will only be implanted once.

### Administration

The following procedures will be performed by the surgeon for administration of the intervention.

1. A clinical examination under anesthesia with an anal fistula probe will be conducted to rule out active infection, anal stenosis, or active proctitis and confirm the presence of a single fistula tract.
2. The seton will be removed, the fistula tract curetted vigorously to remove granulation tissue lining the tract, and the tract will be flushed with saline.
3. The fistula tract length and diameter and amount of the sphincter muscle involved will be assessed by the surgeon with an anal fistula probe placed from the external to the internal opening.
4. The appropriate model collagen matrix will be used based on the length of the tract.
5. The collagen will be saturated with the rhPDGF solution in a sterile dish for a minimum of 10 min.
6. After the collagen is fully saturated with rhPDGF, it will be pulled through the fistula tract carefully from the internal opening to the external opening.
7. The collagen will be sutured to the rectal mucosa at the internal opening and any portion beyond the skin level will be trimmed to sit flush with the external opening.

### Outcomes

#### Primary outcome

The primary outcome of this study will be to evaluate the technical performance of rhPDGF-enhanced collagen implantation for treatment of complex perianal fistulas. This will be measured by the proportion of participants with procedural success as defined by implantation of the intervention without any intervention-related serious adverse events (SAEs) within 3 months of the procedure. The 3-month endpoint is the average timepoint when the surgeon makes the diagnosis that the perianal fistula has completely healed or failed to heal.

#### Secondary outcomes

The secondary outcomes of this study will be used to assess the preliminary safety and efficacy of rhPDGF-enhanced collagen for treatment of complex perianal fistulas. A quantitative and descriptive summary of all intervention-related adverse events (AEs) within 3 months of the procedure will be used to assess preliminary safety. Clinical evaluation of fistula healing assessed as healed/not healed by no drainage/drainage from the external opening at the Month 3 follow-up visit will determine the proportion of fistulas healed. Rate of recurrence is assessed by the number of participants with clinically diagnosed healed fistulas (no drainage/no leakage) that are later diagnosed with recurrence at the final, Month 6 study visit. The Month 6 endpoint was chosen because successful treatment of complex perianal fistulas is based not only on a diagnosis of being completely healed by clinical exam but also on clinical remission without recurrence, and the majority of recurrences occur within 6 months of the treatment. Safety and efficacy will also be assessed by the change in patient-reported pain, incontinence, and other perianal fistula-related symptoms in the baseline and weekly follow-up participant surveys.

#### Tertiary outcomes

The tertiary outcomes of this study will assess the feasibility of studying rhPDGF-enhanced collagen for treatment of complex perianal fistulas in a larger efficacy trial. This will be determined by total number of participants recruited and eligible, number of participants randomized per month, and the proportion of participants retained at Month 3 and Month 6.

#### Crossovers

If the fistula is not healed at the Month 3 study visit and the participant is in the control arm of the study, they will reach their endpoint as a control and have the option to crossover to the investigational arm and receive the rhPDGF-enhanced collagen treatment. These participants will first have another draining seton placed in their fistula tract for approximately 4 weeks and be scheduled for implantation of the rhPDGF-enhanced collagen. This group will restart the schedule of activities at visit 2 for the procedure and return for all post-procedural study visits 3-5 at Months 1, 3, and 6. Their outcome measures will be tracked and documented, and they will complete the weekly participant surveys, but their results will not be included in the initial primary analyses of the original randomized participant data. This will be considered extended, open-label use of the investigational intervention and will not delay reporting the outcomes from the comparison of healing between the original intervention and control groups at Month 3. Their outcome measures will be analyzed and reported later.

### Study Assessments and Procedures

#### Safety Assessments

- Clinical examination

- At screening, during the procedure, and at all follow-up visits, clinical examination of the perianal fistula will visually assess for infection, excess drainage, chronic inflammation, abnormal bleeding, or fibrosis at the external opening. Photographs of the external openings will be taken at screening and all follow-up visits to document healing progress.
- Patient medical history

- At screening, patients meeting inclusion criteria will be screened for exclusion criteria through their medical history. The etiology of the perianal fistula will also be discussed and documented. Participants will review any known allergies, previous surgical procedures, previous complications, and frequency of fistula occurrence or recurrence.
- Pregnancy test

- At screening, women of childbearing potential will be required to take a pregnancy test and must agree to use two reliable contraceptive methods for the duration of their participation in the study, one of which must be a barrier method; the contraceptive methods will be documented in the screening eCRFs.
- Crohn’s Concomitant Medications

- At screening and all follow-up visits, Crohn’s medications will be documented for patients with CD-related perianal fistulas, including medication name, single dose amount, route, frequency, start date, and end date.
- Participant survey

- At screening, baseline, and weekly following the procedure till Month 3, participants will complete a self-report questionnaire assessing the frequency and severity of pain, incontinence, and other perianal fistula-related symptoms. If symptoms are increasing in frequency and severity, an off-cycle clinical visit will be scheduled to address any adverse events.
- Assessment of adverse events

- The following adverse events will be recorded in the eCRFs and followed until resolution:

- Adverse events that are classified as related to study procedures (that is, potentially-, probably-, or definitely related to study procedures or of uncertain relationship to study procedures), regardless of severity
- Serious adverse events regardless of relatedness to study procedures.
- Clinical adverse events, events diagnosed as clinical conditions, with a severity grade ≥ 3, regardless of relatedness to study procedures.

#### Efficacy Assessments

- Clinical examination

- At screening, baseline, and all follow-up visits, clinical examination of the perianal fistula will visually assess drainage, local inflammation, and epithelialization at the external opening to determine if the fistula is healed. Photographs of the external openings will be taken at screening and all follow-up visits to document healing progress.
- Baseline Participant Survey

- At consenting, participants will complete a self-report questionnaire to assess:

- Perianal fistula-related symptoms
- Pain scale
- Wexner Incontinence Scale
- Follow Up Participant Survey

- Following the baseline procedure till the Month 3 follow-up visit, participants will complete a weekly self-report questionnaire to assess:

- Perianal fistula-related symptoms
- Pain scale
- Incontinence (adapted from Wexner Incontinence Scale)
- Study Intervention – Baseline or Crossover

- Seton removal, debridement of fistula tract, and implantation of the rhPDGF-BB-enhanced collagen plug
- Study Control – Baseline only

- Seton removal and debridement of fistula tract – Routine care for CD-related complex perianal fistulas
- Seton removal, debridement of fistula tract, and implantation of the commercially available anal fistula plug – Routine care for idiopathic complex perianal fistulas

### Participant Discontinuation/Withdrawal from the Study

Participants are free to withdraw from participation in the study at any time upon request. An investigator may discontinue or withdraw a participant from the study for the following reasons:

- Failure to heal by the Month 3 follow-up visit
- Infection or abscess which requires discontinuation of the study intervention by removal of the collagen plug followed by a routine care procedure
- Unacceptable adverse event(s)
- In the judgement of the investigator, further treatment would not be in best interest of patient
- Substantial non-compliance by the patient with the requirements of the study
- The patient uses illicit drugs that may have reasonable chance of interfering with results
- Patient is lost to follow-up
- Development of intercurrent illness or situation which would affect assessments of clinical status and study endpoints to a significant degree
- Pregnancy

- In the event a study participant becomes pregnant prior to the Month 3 follow-up visit, the pregnancy will be followed to delivery and maternal and fetal outcomes will be documented.
- Incarceration
- Disease progression which requires discontinuation of the study intervention
- Participant meets an exclusion criterion (either newly developed or not previously recognized) that precludes further study participation

### Risks

The inclusion and exclusion criteria for this study were designed to minimize the risks to participants receiving rhPDGF-enhanced collagen for treatment of complex perianal fistulas in this study. Given that the intervention will be implanted at baseline and the rhPDGF will be released and metabolized over the first 2-3 days, we do not anticipate any long-term risks. The plethora of human safety data that is published supports the expectation that the only risks involved with the use of rhPDGF is the potential to increase acute inflammation at the site of application following intervention. There is the possibility of erythematous at the site of application due to the accumulation of blood in dilated capillaries during the initial healing phases, but these are also the primary symptoms associated with the presence of an anal fistula. The only other risks are associated with routine care procedures. The use of a commercially available collagen plug is one type of routine care procedure with a success rate of approximately 50%. The collagen matrix used in the intervention arm of the study will be saturated with rhPDGF to potentially enhance healing and improve the success rate. Throughout the life cycle of the study, safety assessments will be used to ensure proper monitoring of adverse events. Safety assessments will consist of clinical examinations, evaluation of patients’ medical history, pregnancy testing, identification of concomitant medication taken due to CD, and weekly participant surveys. The potential benefits of this investigational intervention to improve the healing rates, time in remission, and quality of life outweigh the minimal risks in individuals with complex perianal fistulas.

### Statistical analysis

Categorical and binary data will be presented using counts and percentages. Continuous data will be presented using medians and interquartile ranges. All statistical tests will use an α = 0.05 Type I error rate, using two-tailed p-values as appropriate for the specified hypotheses, but 95% confidence intervals will be given primary focus above p-values (29). Statistical models will be adjusted for Crohn’s status. Model assumptions will be checked using visualizations and tests as appropriate, with data transformations and alternative models considered when assumptions are dubious.

#### Sample Size

This study will aim to recruit and screen 60 eligible patients to enroll and randomize 12 participants in a 2:1 ratio. Sample size was influenced by expected confidence interval widths for the primary outcome (technical performance of the rhPDGF-BB-enhanced collagen implant for treatment of complex perianal fistulas). Assuming that all 8 participants assigned to the intervention arm will achieve successful implementation of the treatment, the lower bound of a Clopper-Pearson 95% confidence interval would be 0.63, indicating high confidence that the treatment does not have a low probability of successful implementation. Sample size was also influenced by the efficacy outcome of healing at 3 months. Based on the prior work in the literature, we assume that the routine care arm (across the expected distribution of routine care procedures) will achieve 50% healed at 3 months (2/4 healed). If 7 of the 8 intervention arm patients achieve healing, the expected lower bound of a Carlin-Louis 95% confidence interval for the differences in proportions healed would be about –0.14, indicating high confidence that the intervention does not cause substantial harm. Furthermore, if all 8 intervention arm patients achieve healing, the expected interval lower bound will be –0.01, such that there is high confidence of treatment benefit.

We assumed that study dropout before Month 3 will not occur, and that crossover into the intervention arm will not occur until after ascertainment of Month 3 outcomes. The time-to-healing will be considered for secondary analyses and likely have slightly higher power assuming that patient-reported data is accurate and high quality, as it achieves higher granularity to assess healing than the binary outcome of healing at Month 3 follow-up. Likewise, exploratory analyses including crossover patients may improve power for assessing the primary outcome. Other secondary endpoints and subgroup analyses are exploratory in nature, and we did not perform formal power analyses for these.

#### Primary Outcomes Analysis

The primary endpoint is technical performance of the treatment. This will be reported as the proportion of participants assigned to the treatment arm with successful implementation of the treatment (without SAEs). This will be reported along with a Carlin-Louis 95% confidence interval.

#### Secondary and Tertiary Outcomes Analysis

The secondary efficacy and safety endpoints will be analyzed for all randomized participants, as appropriate. Analyses will not be dependent on the findings of the primary endpoint. The first secondary endpoint considers all treatment-related adverse events and the proportion of participants with such events by arm will be analyzed/reported in the same way as the primary endpoint. The proportion will then be compared across arms using Fisher’s Exact test. If there are at least two participants with multiple treatment-related adverse events, a Mann-Whitney test will compare the number of adverse events per participant across arms.

The next secondary endpoint is fistula healing at Month 3 measured as a binary variable. The probability of healing in each arm will be calculated as the proportion of randomized participants per arm that are diagnosed as completely healed by a clinical evaluation of no visible drainage from the external opening at the Month 3 follow-up visit. The endpoint will be modeled as the outcome in a logistic regression model, conditioning on the treatment arm and baseline etiology. The effect will be presented as an adjusted odds ratio with a 95% confidence interval, as well as the marginal risk ratio after performing standardization. A p-value less than 0.05 will be considered statistically significant, but interpretation will be primarily focused on the 95% confidence interval. Residuals will be inspected to inspect model fit and validity of the homoscedasticity assumption. If model fit is poor, a Fisher’s Exact test will be used in place of logistic regression. Missing data are expected to be limited but will be handled using multiple imputation. Individuals who are lost to follow-up before Month 3 will also be included via the multiple imputation procedure, considering plausible imputations conditional on their baseline and Month 1 follow-up assessments, as well as weekly symptom data. The primary analysis will be modified intention-to-treat (ITT) based on randomized arm, as the investigational product is not available to patients who are not randomized to treatment before the Month 3 outcome assessment. It will be modified ITT in the sense that only participants who receive the investigational intervention will be analyzed as part of the intervention arm; this is necessary as some exclusion criteria cannot be assessed until the time of surgery.

Supplementary analysis of this secondary endpoint will consider time in weeks to achieve complete healing from baseline up to 12 weeks. This endpoint will be determined by the self-reported measure of no leakage from the weekly participant survey and will be focused on the first achievement of complete healing, as recurrence will be assessed in other outcomes. This endpoint will be analyzed using a Cox proportional hazards model, conditional on the treatment arm and Crohn’s status as outlined in the SAP. The results will be presented as an adjusted hazard ratio and a marginal hazard ratio after performing standardization. Proportional hazards will not be checked with a hypothesis test, as this is typically not a recommended procedure; instead, Kaplan-Meier curves will be calculated, and plausibility of proportional hazards will be assessed visually. If proportional hazards are violated or model fit is poor, the outcome will be compared across arms using the log-rank test. Missing data are expected to be limited, but will be handled using multiple imputation, taking the longitudinal nature of surveys into account.

The next secondary endpoint is fistula recurrence at 6 months. This will be measured as the percentage of the total number of treated participants in each arm that were diagnosed as healed at Month 3 with subsequent recurrence of the fistula by clinical evaluation of the external opening at the Month 6 follow-up visit. This is a binary endpoint which will be analyzed using the same procedures as those outlined for the binary Month 3 healing endpoint.

The final secondary endpoint is the change in the severity of symptoms at Month 3 (adjusted for baseline), which will be calculated using the ordinal scale for each question in the weekly participant survey. Scores will be compared between arms using Mann-Whitney tests. A composite variable will be calculated as defined in the SAP and analyzed using ordinal logistic regression conditional on treatment arm, baseline score, and baseline etiology. The proportional odds assumption will be assessed using a parallel line plot. A longitudinal ordinal mixed effects model with similar specification and a random intercept for study participant and a linear time term will be used to compare longitudinal symptom severity over time. Results will be presented as adjusted odds ratios and missing data will be handled using multiple imputation.

Tertiary outcomes will be reported as totals or proportions, as appropriate. Analyses of proportions retained at 3 months and 6 months will be similar to those for the primary outcome. Exploratory analyses which consider outcomes of participants who cross over into the investigational device arm will be reported separately or included in sensitivity analyses.

### Trial status

Screening, enrollment, and randomization for this trial began in November 2024. Data collection will be completed between August – December 2026 depending on crossover status and results will be reported shortly after. The trial is still ongoing at the time of submission.

### Patient and public involvement

Patients and public were not involved in the design, conduct, reporting, or dissemination plans for this research.

## ETHICS AND DISSEMINATION

Participant confidentiality and privacy is strictly held in trust by the study team, and all research activities will be conducted in as private a setting as possible. This confidentiality is extended to cover the participant’s clinical information and the study protocol, documentation, data, and all other information generated. No information concerning the study or data will be released to any unauthorized third party. All faculty and staff working on the study receive and provide documentation of training in Good Clinical Practice (GCP) as part of their onboarding and continuing education.

Data, safety, and enrollment monitoring is conducted to ensure that the rights and well-being of trial participants are protected, that the reported trial data are accurate, complete, and verifiable, and that the conduct of the trial is in compliance with the currently approved protocol/amendment(s), with International Conference on Harmonisation Good Clinical Practice (ICH GCP), and with applicable regulatory requirement(s). Safety oversight will be under the direction of a Data and Safety Monitoring Board (DSMB) composed of 3 independent members. They will be experts/representative: clinicians, clinical trialists, and statisticians. They will not be study investigators or involved in execution of the study protocol. Furthermore, no member should have financial, proprietary, professional, or other interests that may affect impartial, independent decision-making by the DSMB.

This study will comply with the NIH Data Sharing Policy and Policy on the Dissemination of NIH-Funded Clinical Trial Information and the Clinical Trials Registration and Results Information Submission rule. As such, this trial will be registered at ClinicalTrials.gov, and results information from this trial will be submitted to ClinicalTrials.gov. In addition, every attempt will be made to publish results in peer-reviewed journals.

## DISCUSSION

In this trial, we used a novel drug repurposing approach, leveraging PheWAS, to identify a novel pathophysiological phenotype that may benefit from a targeted pharmacological intervention (1–3). Utilizing VUMC’s Synthetic Derivative (SD), a database of almost 4 million de-identified electronic health records, and BioVU, a biobank of over 300,000 deidentified patient DNA samples with SNV genotyping data, PheWAS revealed that patients carrying SNV rs41287112 in *PDGFRB,* causing decreased PDGFRβ protein expression, have a significantly higher prevalence of chronic wounds and tissue defects, including fistulas. PDGF binds PDGFRβ and is the key activator and regulator of biological mechanisms that drive innate healing, and recombinant human PDGF (rhPDGF) mimics the endogenous protein and is FDA-approved for healing cutaneous wounds and periodontal and orthopedic tissue regeneration. We hypothesize that rhPDGF will enhance healing and remission of complex perianal fistulas by precisely targeting the mechanisms of tissue repair.

This hypothesis is further supported by studies demonstrating improved healing rates for many types of fistulas, including perianal fistulas, with platelet rich fibrin (PRF) or platelet-rich plasma (PRP) (22–28), and it is accepted that PDGF is the main driver of these benefits due to its high concentrations in platelets. However, the concentration and effectiveness of these blood-derived products is highly variable based on the patient’s age and health and the blood preparation technique. In humans, topical rhPDGF was used to heal post-laryngectomy fistulas, and the data showed that daily treatments stimulated significant granulation tissue formation and a 50% reduction in the fistula size after one week (30). Collectively, the previous scientific evidence showing exogenous PDGF enhances healing in various types of fistulas and novel PheWAS data which implicates impaired PDGF signaling in chronic wound development suggest that rhPDGF has promise for providing therapeutic benefits in complex perianal fistulas, which remain a major clinical challenge in need of new multimodal treatments aimed at tissue repair and regeneration.

This prospective, double-blind, randomized-controlled clinical trial aims to evaluate the safety and efficacy of rhPDGF-enhanced collagen for treatment of complex perianal fistulas. The natural history and actual case-counts of the two patient populations, idiopathic and CD-related fistulas, guided the trial design, objectives, outcomes measures, and sample size with a goal of completing enrollment in approximately 15 months. A 2:1, intervention versus control, randomized parallel arm design was chosen for the purposes of evaluating preliminary performance, safety, efficacy, and feasibility. The control procedures were selected based on the common routine care options for each patient population that will offer meaningful comparisons between the two arms. Patient’s that may benefit from a different routine care approach will be excluded from the trial at the surgeon’s discretion. The secondary objectives assessing the potential efficacy of this treatment, such as healing, remission, and change in symptoms, will include stratified subgroup analyses for idiopathic and CD-related fistulas due to the pathophysiology and prognosis differences in these two groups. For this pilot trial, we used both published data and the investigator’s real-world experience with healing in these patients to design the objectives and outcomes that will guide a larger more definitive follow up trial where assessing efficacy may or may not include both etiologies. Additionally, some patients have lived with persistent fistulas for years after conventional treatments have failed making these patients harder to recruit for a study that may ultimately result in another unsuccessful attempt. To enhance patient recruitment and retention, a crossover design was employed, allowing patients within the control group the opportunity to receive the intervention if their fistula fails to heal by the Month 3 follow-up visit.

Several challenges and limitations with this clinical study must be acknowledged. Most anal fistulas are simple and can be successfully treated by fistulotomy, which is the well-established gold standard with 90-95% success rates (31). Complex perianal fistulas are rare and present with diverse anatomical characteristics, which, under routine care, require a creative approach to optimal treatment selection by the surgeon, and some would not benefit from an implantable scaffold simply due to the physical shape or branching of the tract. Stringent exclusion criteria were required to establish a study population who might benefit the most from rhPDGF-enhanced collagen treatment or the control procedures under routine care, but the small sample size restricts the ability to perform subgroup analyses and limits statistical power for efficacy outcomes. Furthermore, the control procedures differ based on disease etiology, with idiopathic patients receiving an FDA approved anal fistula plug while the CD patients are left to heal naturally, yet all patients undergoing treatment with rhPDGF, regardless of disease etiology, will also receive the implanted collagen matrix. As such, the interpretation of the secondary outcomes may be difficult due to the lack of a true placebo. Despite these limitations, if successful, this trial would guide a larger, multi-site efficacy trial for a much-needed alternative treatment option for complex perianal fistulas and support the utility of using PheWAS for drug development and repurposing.

## Data Availability

All data produced in the present study are available upon reasonable request to the authors

## AUTHORS’ CONTRIBUTIONS

Study concept and design: ATH, JKS, JMP, SEL, CRC, and BSB. Revisions and refinements of study protocol: ATH, JS, JKS, JMP, SEL, AK, CRC, and BSB. Acquisition and management of data: MCK, SAY, MDL, ATH, JS, TEU, and AK. Drafting and writing of the manuscript: MDL, CRC, and BSB. Manuscript review and revisions: MCK, SAY, MDL, ATH, JS, JKS, JMP, SEL, TEU, AK, CRC, and BSB. Study supervision: ATH and CRC.

