## Supplementary material for "Study protocol and statistical analysis plan for a randomized controlled trial evaluating the safety and feasibility of the recombinant human platelet-derived growth factor B (rhPDGF-BB)-enhanced collagen plug for complex perianal fistula healing": S2 Appendix

**IND Sponsor:** Vanderbilt University Medical Center
1211 21$^{st}$ Avenue South
Nashville, TN 37212

**Funder:** Lynch Regenerative Medicine

**ClinicalTrials.gov number:** NCT06632418

**IRB number:** 240585

**Protocol Version Number:** V.6.1

**Protocol Date:** 13 December 2024

**Study Team Leadership**

| | |
|---|---|
| Principal Investigator | Alexander T. Hawkins, MD, MPH<br>Director, Colorectal Research Center<br>Associate Professor of Surgery<br>Vanderbilt University Medical Center |
| Primary Statistician | Bryan Blette, PhD<br>Assistant Professor of Biostatistics<br>Vanderbilt University Medical Center |
| Project Manager | Cyndi R. Clark, PhD<br>Senior Scientific Research Project Manager<br>Vanderbilt Institute for Clinical & Translational Research<br>Vanderbilt University Medical Center |
| Project Director | Jill M. Pulley, MBA<br>Research Professor of Medicine<br>Vanderbilt Institute for Clinical & Translational Research<br>Vanderbilt University Medical Center |
| Coordinating Center Lead | Wesley H. Self, MD, MPH<br>Senior Vice President for Clinical Research<br>Professor of Emergency Medicine<br>Vanderbilt Institute for Clinical & Translational Research<br>Vanderbilt University Medical Center |

### 1 INVESTIGATIONAL PLAN

The trial will be carried out in accordance with International Conference on Harmonisation Good Clinical Practice (ICH GCP) and the following:

- United States (US) Code of Federal Regulations (CFR) applicable to clinical studies (45 CFR Part 46, 21 CFR Part 50, 21 CFR Part 56, 21 CFR Part 312, and/or 21 CFR Part 812)

The protocol, informed consent form(s), recruitment materials, and all participant materials will be submitted to the Institutional Review Board (IRB) for review and approval. Approval of both the protocol and the consent form must be obtained before any participant is enrolled. Any amendment to the protocol will require review and approval by the IRB before the changes are implemented to the study. In addition, all changes to the consent form will be IRB-approved; a determination will be made regarding whether a new consent needs to be obtained from participants who provided consent, using a previously approved consent form.

#### 1.1 SYNOPSIS

| | |
|---|---|
| **Title:** | A randomized controlled trial evaluating the safety and feasibility of the recombinant human platelet-derived growth factor B (rhPDGF-BB)-enhanced collagen plug for complex perianal fistula healing |
| **Short Title:** | rhPDGF-BB-enhanced collagen plug for perianal fistula healing |
| **Principal Investigator:** | Alexander Hawkins, MD, MPH |
| **Investigational Intervention Name:** | Recombinant human platelet-derived growth factor B **(**rhPDGF-BB)-enhanced collagen plug |
| **Intervention Description:** | 510k dual surface collagen membrane hydrated with rhPDGF-BB solution |
| **Study Description:** | An early phase IIa study to evaluate the technical performance, preliminary safety and efficacy, and feasibility of the recombinant human platelet-derived growth factor B (rhPDGF-BB)-enhanced collagen plug for healing complex perianal fistulas not amenable to fistulotomy |
| **Rationale:** | Complex perianal fistulas are not eligible for a fistulotomy due to the risk of bowel incontinence. Routine care for these patients is draining seton placement followed by seton removal and natural healing, endorectal advancement flap, LIFT procedure, anal fistula plug, or some combination of procedures; however, the average success rate for healing and remission of complex perianal fistulas is approximately 50%. A phenome-wide association study (PheWAS) suggested that patients with a single nucleotide variant (SNV) in PDGFRβ have a higher incidence of fistulas, leading to the hypothesis that the loss of PDGFβ signaling impairs tissue integrity and regrowth after injury, infection, or inflammation. rhPDGF-BB, a recombinant human platelet derived growth factor protein-based therapy, signals through PDGFRβ to recruit fibroblasts and pericytes and promote angiogenesis and epithelialization in soft tissue and is used to enhance bone and soft tissue growth in patients with periodontitis. Building |

-- Confidential --
This document intended for restricted use only and may not – in full or part – be passed on, reproduced, published, or used without express permission.
All rights reserved.

| | |
|---|---|
| | on these prior data, this clinical trial will evaluate the technical performance, preliminary safety and efficacy, and feasibility of the rhPDGF-BB-enhanced collagen therapy on soft tissue regrowth and stability for the treatment of complex perianal fistulas. |
| **Objectives:** | **Primary Objective:** To evaluate the technical performance of the rhPDGF-BB-enhanced collagen plug for treatment of complex perianal fistulas<br>**Secondary Objectives:** To assess the preliminary safety and efficacy of the rhPDGF-BB-enhanced collagen plug for treatment of complex perianal fistulas<br>**Tertiary Objectives:** To determine the feasibility of studying the rhPDGF-BB-enhanced collagen plug for treatment of complex perianal fistulas in a definitive trial |
| **Outcome Measures:** | **Primary Outcome Measure:** The proportion of participants with procedural success as defined by implantation of the device without device-related serious adverse events (SAEs) within 3 months of the procedure<br>**Secondary Outcome Measures**:<br>1. Quantitative and descriptive summary of all device-related adverse events (AEs) within 3 months of the procedure<br>2. Proportion of fistulas healed at 3 months<br>3. Recurrence of fistulas at 6 months<br>4. Baseline and Weekly Follow-Up Participant Surveys (for 3 months following the procedure)<br>a. Change in symptoms<br>b. Change in pain<br>c. Change in incontinence score (adapted from Wexner Incontinence Score)<br>**Tertiary Outcomes:**<br>1. Total number of participants recruited and eligible<br>2. Number of participants randomized per month<br>3. Proportion of participants retained at 3 months and 6 months |
| **Study Population:** | **Key Inclusion Criteria:**<br>- Diagnosis of a single tract perianal fistula not amenable to fistulotomy as determined by the supervising surgeon<br>- Fistula must currently have a draining seton in place<br>- Aged >21 years old<br>- Willing and able to provide informed consent and to comply with study protocol and follow-up<br>- Stated willingness to comply with all study procedures and availability for the duration of the study<br><br>**Key Exclusion Criteria:**<br>- Medical conditions that would, in the opinion of the investigator or treating provider, compromise the safety of the individual with study participation and/or the ability of the individual to follow study protocol<br>- Genito-urinary fistulization, including rectovaginal (i.e., fistulas that transverse the vaginal canal)<br>- Presence of an ileal anal pouch<br>- Any major surgery of the gastrointestinal tract (including one or more segments of the colon or terminal ileum) within 3 months prior to randomization; presence of stoma is not exclusionary |

-- Confidential --
This document intended for restricted use only and may not – in full or part – be passed on, reproduced, published, or used without express permission.
All rights reserved.

| | |
|---|---|
| | - Prior surgical procedure (i.e., Ligation of Intersphincteric Fistula Tract or Endorectal Advancement Flap) for the target fistula or a perianal procedure that resulted in a large soft tissue defect within 6 months prior to screening visit<br>- One or more of the following fistula types or anatomic presentations: horseshoe fistulas, fistulas that do not have an opening inside the anal canal or low rectum, blind ending sinus tracts (no external opening), branching fistulas (a previously performed conversion of a branching fistula tract to a single tract is not exclusionary), >1 internal opening, moderate or severe proctitis, severe rectal mucosal fibrosis surrounding the internal opening preventing the securing of the fistula plug disk, any anatomical limitation to successfully securing the fistula plug disk.<br>- Known allergic reactions to porcine collagen or yeast-derived products<br>- Currently enrolled in a drug or device trial or within 30 days of last investigational drug or device administration at baseline visit where investigational treatment (drug or device) was placed in or near the fistula tract or may potentially interact with study treatment<br>- Women who are pregnant, breastfeeding, or planning to become pregnant during the trial<br>- Active infection at the application site<br>- The presence of malignant neoplasms at the application site<br>- Prior radiation therapy at the application site |
| **Phase:** | Phase 2a Clinical Trial |
| **Description of Sites/Facilities Enrolling Participants:** | Vanderbilt University Medical Center (VUMC) |
| **Description of Fistula Treatment in Intervention Group:** | 510k porcine-sourced collagen membrane saturated with rhPDGF-BB, supplied as GEM 21S, consisting of pure, sterile 0.3 mg/mL rhPDGF-BB in 20 mM sodium acetate buffer pH6+/-0.5 from a pre-filled syringe |
| **Description of Fistula Treatment in Control Group:** | Controls will receive routine care for Crohn's or idiopathic perianal fistulas. Routine care includes seton removal followed by natural healing for Crohn's or implantation of a commercially available anal fistula plug for idiopathic. |
| **Study Duration:** | 15 months |
| **Participant Duration:** | 6 months |
| **Blinding:** | Participants and follow-up assessors will be blinded |
| **Sample size:** | 12 |
| **Randomization** | 2:1 permuted randomization with blocks of size 3 |
| **Stratification:** | Blocked randomization stratified by Crohn's:idiopathic etiology |

-- Confidential --
This document intended for restricted use only and may not – in full or part – be passed on, reproduced, published, or used without express permission.
All rights reserved.

#### 1.1.1 SCHEMA

**Enroll**: Obtain informed consent. Screen potential participants by inclusion and exclusion criteria; obtain history, complete baseline participant survey, document.

Randomize
2:1 Intervention:Control

rhPDGF-BB + Collagen Plug
8 participants
Stratification:
Crohn's : Idiopathic

Routine Care
4 participants
Stratification:
Crohn's : Idiopathic

**Intervention Procedure**

1. A clinical examination under anesthesia will be conducted to rule out active infection, anal stenosis, or active proctitis.
2. The seton will be removed, the fistula tract curetted vigorously to remove granulation tissue lining the tract, and the tract will be flushed with saline.
3. The fistula tract length and diameter and amount of sphincter muscle involved will be assessed by the surgeon with an anal fistula probe
4. The appropriate model collagen membrane will be used based on the length of the tract.
5. The collagen membrane will be cut into strips that are longer than the fistula tract to provide enough material for suturing and trimming.
6. The strips will be collected and secured by a loop to create the fistula plug
7. The plug will be saturated with the rhPDGF-BB solution in a sterile dish for at least 10 min
8. After the plug is fully saturated with rhPDGF-BB, it will be pulled through the fistula tract carefully from the internal opening to the external opening
9. The collagen plug will be sutured to the rectal mucosa at the internal opening and any portion beyond the skin level will be trimmed to sit flush with the external opening.
10. Participant will be discharged with at-home instructions and follow-up visit schedule

**Control Procedure**

1. A clinical examination under anesthesia will be conducted to rule out active infection, anal stenosis, or active proctitis.
2. The seton will be removed, the fistula tract curetted vigorously to remove granulation tissue lining the tract, and the tract will be flushed with saline.
3. The fistula tract length and diameter and amount of sphincter muscle involved will be assessed by the surgeon with an anal fistula probe
4. The physician will perform the routine care procedure based on the etiology of the fistula
   a. Natural healing – Crohn's
   b. Commercially available anal fistula plug - idiopathic
5. Participant will be discharged with at-home instructions and follow-up visit schedule

**At home**
Weekly for 3 months

**Follow-up assessments of study endpoints and safety:**

1. Follow Up Participant Survey (REDCap)

**Visit 3-4**
Months 1, 3

**Follow-up assessments of study endpoints and safety:**

1. Visual assessment of external opening for drainage
2. Photographic images of external opening
3. Adverse events

**Visit 5**
Month 6

**Final Assessments:**

1. Visual assessment of external opening for recurrence
2. Photographic images of external opening

-- Confidential --
This document intended for restricted use only and may not – in full or part – be passed on, reproduced, published, or used without express permission.
All rights reserved.

##### 1.1.2 SCHEDULE OF ACTIVITIES

| Procedures | Screening Weeks -6 to -4 | Randomized/Baseline Visit 2, Day 0 | At home, Weeks 1-4 Every 7 days +/- 2 days | Study Visit 3 Month 1 +/- 5 days | At home, Weeks 5-12 Every 7 days +/- 2 days | Study Visit 4 Month 3 +/- 5 days | Final Study Visit 5 Month 6 +/- 5 days |
|---|---|---|---|---|---|---|---|
| Informed consent | X | | | | | | |
| Demographics | X | | | | | | |
| Medical history | X | | | | | | |
| Pregnancy test (women of childbearing potential) | X | | | | | | |
| Baseline Participant Survey | X | | | | | | |
| Randomization | X | | | | | | |
| Administer study intervention | | X | | | | | |
| Clinical exam | X | | | X | | X | X |
| Clinical exam under anesthesia | | X | | | | | |
| Anoscopy (SoC, as indicated) | | | | X | | X | |
| Follow Up Participant Survey | | | X | | X | | |
| Adverse Event Review | | | | X | | X | |
| Complete Case Report Forms (CRFs) | X | X | | X | | X | X |

#### 1.2 PURPOSE

##### 1.2.1 ABBREVIATIONS

| | |
|---|---|
| AE | Adverse Event |
| CD | Crohn’s disease |
| CFR | Code of Federal Regulations |
| CONSORT | Consolidated Standards of Reporting Trials |
| CPT | Current Procedural Terminology |
| CRF | Case Report Form |
| DSMB | Data Safety Monitoring Board |
| eCRF | Electronic Case Report Forms |
| EMR | Electronic Medical Records |
| EUA | Exam Under Anesthesia |
| FDA | Food and Drug Administration |

-- Confidential --
This document intended for restricted use only and may not – in full or part – be passed on, reproduced, published, or used without express permission.
All rights reserved.

| | |
|---|---|
| GCP | Good Clinical Practice |
| GEM 21S | Growth-factor Enhanced Matrix 21S |
| GLP | Good Laboratory Practices |
| HIPAA | Health Insurance Portability and Accountability Act |
| IB | Investigator's Brochure |
| ICD-10 | International Classification of Diseases, Tenth Revision |
| ICF | Informed Consent Form |
| ICH | International Conference on Harmonisation |
| IDE | Investigational Device Exemption |
| IFU | Instructions for Use |
| IND | Investigational New Drug |
| IRB | Institutional Review Board |
| ITT | Intention-To-Treat |
| LIFT | Ligation of Intersphincteric Fistula Tract |
| MOP | Manual of Procedures |
| NIH | National Institutes of Health |
| OR | Operating Room |
| PDGFB | Platelet-Derived Growth Factor B |
| PDGFRβ | Platelet-Derived Growth Factor Receptor Beta |
| PheWAS | Phenome-Wide Association Study |
| PI | Principal Investigator |
| PMA | Pre-Market Approval |
| QA | Quality Assurance |
| QC | Quality Control |
| QoL | Quality of Life |
| REDCap | Research Electronic Data Capture |
| rhPDGF-BB | Recombinant human Platelet-derived Growth Factor BB |
| SAE | Serious Adverse Event |
| SoC | Standard of Care |
| SAP | Statistical Analysis Plan |
| SNV | Single Nucleotide Variant |
| SOA | Schedule of Activities |
| SOP | Standard Operating Procedure |
| UP | Unanticipated Problem |
| US | United States |
| VUMC | Vanderbilt University Medical Center |

##### 1.2.2 STUDY RATIONALE

***Perianal Fistula***

Perianal fistula, an abnormal connection between the rectum and perianal epithelium, is a common cause of anorectal burden, accounting for up to 5% of proctological consultations and often responsible for substantial detriment to patient quality of life in terms of pain, fecal incontinence, and psychologic distress.[1] The generally

-- Confidential --
This document intended for restricted use only and may not – in full or part – be passed on, reproduced, published, or used without express permission.
All rights reserved.

accepted pathophysiology includes obstruction of an anal gland followed by chronic infection and epithelization of the abscess drainage tract.[2,3] However, mechanistic clarity regarding proposed pro-inflammatory and infectious factors remains elusive.[4–7] The second most common cause of perianal fistulas is due to Crohn's disease (CD), a chronic inflammatory disease of the gastrointestinal tract. CD-related perianal fistulas are a severe complication of chronic intestinal inflammation, extracellular matrix remodeling, and pathogen-associated molecular patterns resulting in epithelial-to-mesenchymal transition and tissue remodeling that pave the fistula tract.[8]

Perianal fistulas are classified by their relationship to the anal sphincter muscles. The popular classification scheme from Parks, Gordon, and Hardcastle describes the anatomic fistula tract relative to anal sphincter muscles: intersphincteric, transsphincteric, suprasphincteric, extrasphincteric, or superficial.[9] Perianal fistulas may also be categorized according to simple versus complex.[10] Complex anal fistula features include high transsphincteric involving ≥30 percent of the external sphincter, suprasphincteric, and extrasphincteric. Additional characteristics indicative of complexity include pre-existing fecal incontinence or association with Crohn's disease, radiation, or malignancy. Simple fistulas lack any complex features and typically refer to superficial, intersphincteric, and low transsphincteric fistulas that involve <30 percent of the external sphincter.

##### *Current Interventions*

Goals of procedural interventions are to heal the fistula tract and preserve anal sphincter function. For patients with preoperative fecal continence and simple fistulas with minimal sphincteric involvement, fistulotomy is often an effective treatment resulting in healing for over 90% of patients.[11] Options for complex fistulas are broader and may include seton placement followed by removal and natural healing, fistulotomy, fistulectomy, ligation of intersphincteric fistula tract (LIFT), endorectal advancement flap, modified Hanley procedure, or anal fistula plug. The average rate of complete healing, likelihood of recurrence, and risk of bowel incontinence varies from procedure to procedure, and there isn't a one-size-fits-all approach to the management of complex perianal fistulas. The routine care option chosen is guided by the initial physician-patient consult and fistula etiology but ultimately resides on the clinical evaluation and opinion of the surgeon once in the operating room.

Non-invasive medical options are lacking for perianal fistulas.[6,12] Recent investigations have focused on targeting the process of collagen remodeling and epithelial to mesenchymal transitions with mixed evidence regarding efficacy.[13] For example, the use of a bioprosthetic anal fistula plug, comprised of an acellular collagen matrix designed to close the internal fistula opening and provide a scaffold for native tissue growth, initially showed promise. However, follow-up studies have demonstrated healing rates of less than 50%.[3] Still, a prospective study of 30 patients with Crohn's perianal fistulas found that an acellular crosslinked porcine dermal collagen matrix was associated with moderate effectiveness at 12 months follow up. An alternative therapy is fibrin glue, which forms a stable clot and promotes migration of fibroblasts and pluripotent cells to lay down collagen and start the healing process. However, existing studies have not shown benefit in terms of healing rates or recurrence rates relative to traditional interventions.[14] Although lacking clear superiority over surgical techniques, the dearth of existing noninvasive options and continued need for sphincter-sparing modalities has justified the continued search for targeted therapies in complex perianal fistulas.

##### *Drug Repurposing Methods*

-- Confidential --
This document intended for restricted use only and may not – in full or part – be passed on, reproduced, published, or used without express permission.
All rights reserved.

Vanderbilt University Medical Center has the largest academic generic drug repurposing program in the U.S. [15–24] Existing in vitro and preclinical models are poor predictors of human biology. Our discovery methods bypass these limitations by providing target validation in humans before the first dose of a drug is ever given. We identify specific variants in drug target genes and use a PheWAS-driven identification of diseases associated with those genetic variants.[25–27] When genetic variant function is established and a known indication of the drug is represented in the human dataset, we can reasonably conclude that the variant recapitulates drug effects. Unlike computer modeling and database linkages, this work uses actual human diseases captured in the clinical record and markers of their pathophysiology. We use the Synthetic Derivative (SD), a fully de-identified database of medical records, and BioVU, a biobank for plasma, DNA samples, and genotyping data, to perform PheWAS and identify novel associations for future clinical development. Importantly, clinical review of de-identified longitudinal patient charts (detail not available through public databases) of affected individuals reveals specific disease presentations that directly guide precision indications and clinical trial design. The precision indication then directly corresponds to the genetic effect of the variant in the gene that is the target of the drug, rather than the set of (typically broad) symptoms of a given disease. With a precision indication, eligibility criteria and endpoints can be directly matched. We propose that precise indications and medically homogenous patient populations increase likelihood of detecting efficacy signals. With this method we reinvigorate and expand the use of older therapies like rhPDGF-BB. The deliberate focus of our method involves therapeutics with established safety profiles, such that enrollment and clinical use can be guided by what is already known.

For this protocol, PheWAS was used to identify single nucleotide variant (SNV) mutations in the *PDGFRB* gene that decrease PDGFRβ signaling in humans and associate these mutations to conditions in patients. Novel phenotypes with unmet medical needs found in the patient health records can be considered as new indications that may benefit from rhPDGF-BB protein therapy. The PheWAS analysis revealed an association between patients with PDGFRβ SNV rs41287112 and a higher incidence of fistulas, suggesting that the loss of PDGFRβ signaling impairs tissue integrity and regrowth after injury or infection.

##### *Platelet-derived Growth Factor*

*PDGFRB*, located on chromosome 5, encodes for platelet derived growth factor receptor beta (PDGFRβ), a monomer of the cell surface tyrosine kinase receptor that plays an essential role during cranial and cardiac neural crest cell development and the formation of blood vessels in the embryo[28]. PDGF-BB, the platelet derived growth factor B ligand, binds PDGFRβ and promotes the recruitment, proliferation, and differentiation of cells during development, wound healing, and tissue regeneration. Specifically, PDGF-BB directly mediates the inflammatory response, fibroblast and pericyte migration and proliferation, and extracellular matrix production and indirectly facilitates endothelial cell recruitment, neovessel development, and re-epithelialization following injury (**Figure 1**). Recombinant human PDGF-BB (rhPDGF-BB) is a manufactured protein-based therapy that is FDA approved for the treatment of diabetic foot ulcers and periodontal defects and may be beneficial in other pathological conditions with a similar physiological mechanism.

Systematic review of preclinical and clinical evidence showed that platelet rich fibrin or plasma improves the healing rates for perianal fistulas[29,30], Crohn's disease-related perianal fistulas[31], vesicovaginal fistulas[32,33], urethrocutaneous fistulas[34], and oroantral fistulas[35], and there is strong scientific rationale that PDGF signaling

-- Confidential --
This document intended for restricted use only and may not – in full or part – be passed on, reproduced, published, or used without express permission.
All rights reserved.

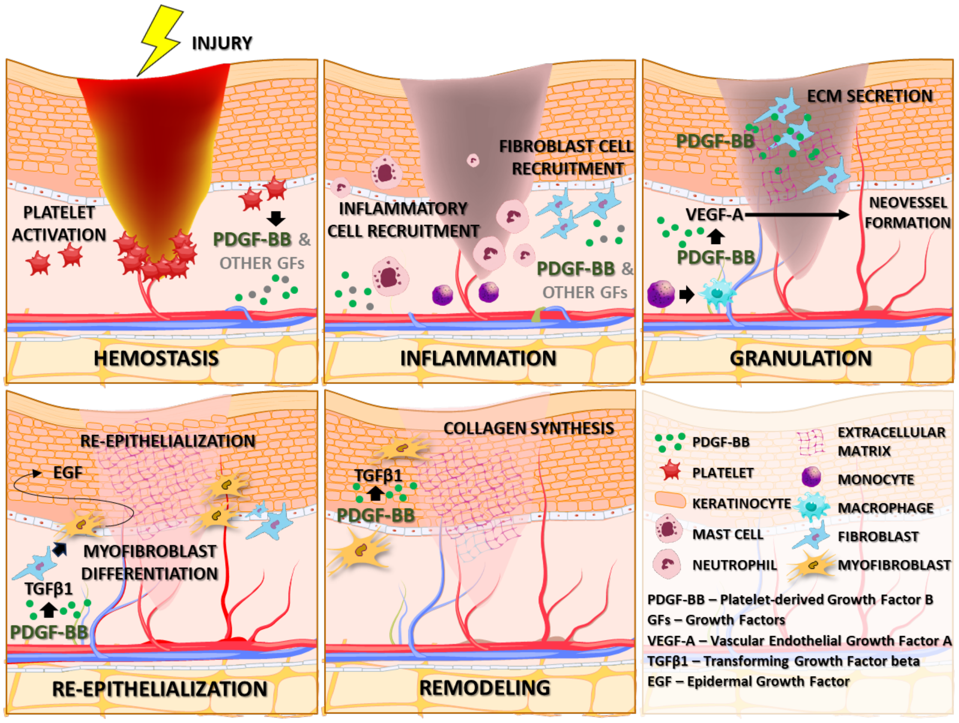

**Figure 1**. PDGF-BB is released by various cell types during all stages of wound healing, binds to PDGFRβ, and signals to mediate inflammatory cell and fibroblasts recruitment (direct), ECM secretion (direct) and neovessel formation (indirect), and re-epithelialization (indirect) and collagen synthesis (indirect).

may drive these benefits due to its high concentrations in platelets and its key role in tissue repair. In humans, topical rhPDGF-BB was used to heal post-laryngectomy fistulas and daily treatments stimulated significant granulation and a 50% reduction in size after one week[36]. Collectively, the scientific evidence reviewed here and novel clinical data from PheWAS suggest that rhPDGF-BB has promise for providing therapeutic benefits in complex perianal fistulas, addressing a major unmet need in the clinic.

***Rationale for evaluating rhPDGF-BB-enhanced dual collagen membrane in perianal fistulas***

Type 1 collagen is a component of the extracellular matrix important to multiple phases of wound healing. Implicated pathways include the induction of fibrin clot formation during hemostasis, recruitment of mesenchymal precursors during the inflammatory phase, and attainment of tensile strength during the maturation phase. Biologic agents comprised of acellular dermal cross-linked collagen such as the Permacol paste (Medtronic) have shown benefit in some studies of Crohn's[37] and idiopathic[38] perianal fistulas, but no consensus exists as other studies found poorer outcomes[39,40]. Monotherapies comprised of individual components of the normal wound healing microenvironment do not capture critical *in vivo* interactions between signaling factors. Preclinical and clinical studies have demonstrated synergistic results of mesenchymal precursors, platelet-rich plasma, and growth factors proteins incubated with bioabsorbable substrate with respect to wound healing.[41] One preclinical study demonstrated the regenerative potential of acellular porcine cartilage enhanced with recombinant PDGF-BB for the reconstruction of septal cartilage defects in a rabbit model[42], and GEM 21S® and Augment® are medical devices comprised of rhPDGF-BB-enhanced matrix indicated for the treatment of periodontal defects and hindfoot and ankle arthrodesis, respectively. More recently, small case series have begun investigating combinatorial therapies for perianal fistulas.[43] Perianal fistulas, idiopathic or CD-related, remain a major clinical challenge in need of new multimodal treatments aimed at tissue repair and regeneration. This protocol aims to evaluate the technical performance, preliminary safety and efficacy, and feasibility for a definitive trial for the rhPDGF-BB-enhanced collagen plug (Geistlich Nexo-Gide® Collagen Membrane) for healing and remission of complex perianal fistulas.

-- Confidential --
This document intended for restricted use only and may not – in full or part – be passed on, reproduced, published, or used without express permission.
All rights reserved.

##### 1.2.3 OBJECTIVES AND OUTCOME MEASURES

| OBJECTIVES | OUTCOME MEASURES | JUSTIFICATION FOR ENDPOINTS |
|---|---|---|
| Primary | | |
| To evaluate the technical performance of the rhPDGF-BB-enhanced collagen plug for treatment of complex perianal fistulas | The proportion of participants with procedural success as defined by implantation of the device without device-related serious adverse events (SAEs) within 3 months of the procedure. | The 3-month endpoint is the average timepoint when the surgeon makes the diagnosis that the treatment for perianal fistula has completely healed or failed . |
| Secondary | | |
| To assess the preliminary safety and efficacy of the rhPDGF-BB-enhanced collagen plug for treatment of complex perianal fistulas | A quantitative and descriptive summary of all device-related adverse events (AEs) within 3 months of the procedure will be used to assess the preliminary safety of this investigational device for the treatment of complex perianal fistulas.<br><br>Clinical evaluation of fistula healing assessed as healed/not healed by no drainage/drainage from the external opening at 3-month follow-up visit will determine the proportion of fistulas healed.<br><br>Rate of recurrence is assessed by the number participants with clinically diagnosed healed fistulas (no drainage/no leakage) that are later diagnosed with recurrence at the final, 6-month study visit.<br><br>Change in symptoms, pain, and incontinence will be assessed by the patient-reported outcomes in the Baseline and Follow Up Participant Surveys completed through Month 3 following the procedure. | The 6-month endpoint was chosen because successful treatment of complex perianal fistulas is based not only on a diagnosis of being completely healed by clinical exam but also on clinical remission without recurrence, and the majority of recurrences occur within 6 months of the treatment. |
| Tertiary | | |
| To determine the feasibility of studying the rhPDGF-BB- | Total number of participants recruited and eligible | Tertiary outcomes are based on the length of the study |

-- Confidential --
This document intended for restricted use only and may not – in full or part – be passed on, reproduced, published, or used without express permission.
All rights reserved.

| OBJECTIVES | OUTCOME MEASURES | JUSTIFICATION FOR ENDPOINTS |
|---|---|---|
| enhanced collagen plug for treatment of complex perianal fistulas in a definitive trial | Number of participants randomized per month<br><br>Proportion of participants retained at 3 months and 6 months | |

#### 1.3 PROTOCOL

##### 1.3.1 OVERALL DESIGN

Perianal fistula is a common colorectal condition with an incidence of 9 cases per 100,000.[44] Most perianal fistulas originate from an anal abscess that does not heal properly and are mainly treated with surgical interventions. The type of surgery and healing rates depend on the classification of the fistula as simple or complex and the proportion of the sphincter muscle that is involved. The second most common cause of perianal fistula is due to Crohn's disease (CD), a chronic inflammatory disease with an estimated annual incidence of 3 to 20 cases per 100,000.[45] Approximately 20% of patients with CD will develop a perianal fistula within 10 years of diagnosis.[46] CD-related perianal fistulas are a severe complication of chronic intestinal inflammation and rely on the combination of pharmacological therapies and surgical interventions, but still one-third of CD-related fistulas remain unhealed.

This Phase II clinical trial will evaluate the performance, preliminary safety and efficacy, and feasibility of the rhPDGF-BB-enhanced collagen plug versus routine care on healing complex perianal fistulas that are not eligible for a fistulotomy. This prospective, blinded, single-site study will randomize participants, 2:1 intervention versus control, into two arms comparing the rhPDGF-BB-enhanced collagen plug to routine care procedures (**Figure 2**) and stratify participants by fistula etiology, Crohn's or idiopathic. Routine care includes draining seton removal and natural healing for Crohn's fistulas or COOK Biotech's Biodesign® Anal Fistula Plug implant for idiopathic fistulas. For both non-CD (idiopathic) and CD-related fistulas, a loose seton is an effective first-line treatment to facilitate drainage of the abscess, prevent the recurrence of an abscess, and increase the chance of healing successfully. After seton placement, the therapeutic interventions for idiopathic and CD-related fistulas differ, and the overall healing rates are widely variable in the literature. The outcomes for CD-related perianal fistulas with seton intervention report success rates between 14-81% [47] and idiopathic perianal fistulas with anal fistula plug intervention are highly similar with success rates between 15.8-72.7% [48].

After recruiting and consenting, participants will be randomized and scheduled for the baseline procedure. Randomization blocks of size 3 (2:1 intervention to control) will be utilized within each stratum (idiopathic versus CD-related), and the patient will be blinded. The investigator will not be blinded because the control and intervention procedures are different. Following the baseline procedure, participants will return for outcomes

-- Confidential --
This document intended for restricted use only and may not – in full or part – be passed on, reproduced, published, or used without express permission.
All rights reserved.

assessment by a blinded physician at one month, three months, and six months and complete a weekly participant survey for 3 months following the procedure from home. At each study visit, the blinded physician will perform a clinical examination of the external opening to assess for inflammation, drainage, and epithelialization, take clinical photographs, and discuss any adverse events (1-month and 3-month visits only). At the 3-month study visit, the blinded physician will determine if the fistula has completely healed, and this will be documented in the study database. At the 6-month final study visit, the blinded physician will assess for recurrence of the fistula by examination of the external opening, and this will be documented in the study database.

If the fistula is not healed at the 3-month study visit and the participant is in the control arm of the study, they will reach their endpoint as a control and have the option to crossover to the investigational arm and receive the investigational intervention. These participants will first have another draining seton placed in their fistula tract for approximately 4 weeks and be scheduled for the intervention procedure, implantation of the rhPDGF-BB-enhanced collagen plug. This group will restart at visit 2 for the procedure and return for all post-procedural study visits 3-5 at 1-, 3-, and 6-months following the procedure (**Figure 2**). Their outcome measures will be tracked and documented, and they will complete the weekly participant surveys, but their results will not be included in the initial analyses of the original randomized participant data. This will be considered extended open-label use of the investigational intervention and will not delay reporting the outcomes from the comparison of healing between the original intervention and control groups at 3 months. Their outcome measures will be analyzed and reported separately.

It is expected that all participants will complete the study which begins at time of signing informed consent and ends at the final study follow-up visit. An overview of the study design is presented in **Figure 2**.

###### 1.3.1.1 SCIENTIFIC RATIONALE FOR STUDY DESIGN

The natural history and actual case-counts of these two patient populations, idiopathic and CD-related complex perianal fistulas, guided the design of the objectives, outcomes, and sample size that would allow this pilot study to be completed in approximately 15 months. A small, 2:1 randomized parallel arm design was chosen for the purposes of evaluating preliminary performance, safety, efficacy, and feasibility. The primary objective of this study is to evaluate the technical performance of the rhPDGF-BB-enhanced collagen plug for the treatment of complex perianal fistulas regardless of etiology. This outcome measure is defined as procedural success of the rhPDGF-enhanced collagen plug without any intervention-related serious adverse events. Procedural complications include but are not limited to acute infection, dislodgement or extrusion of the plug, and non-healing of the fistula tract. The secondary objectives that are focused on the potential efficacy of this treatment will include stratified subgroup analyses for idiopathic and CD-related fistulas due to the differences in natural history and potential differences in outcomes with routine care in these two groups. Further, the stratified randomized design will promote balance of the etiologies across the trial arms. For this pilot trial, we used both published data and the investigator's real-world experience with healing in these patients to design the objectives and outcomes that will guide a larger more definitive follow up trial where assessing efficacy may or may not include both etiologies. Given the number of different perianal fistula types, anatomical presentations,

-- Confidential --
This document intended for restricted use only and may not – in full or part – be passed on, reproduced, published, or used without express permission.
All rights reserved.

and approaches to treatment, the inclusion criteria are specific to fistulas that are a single continuous tract, have had a draining seton placed, and are not amenable to fistulotomy regardless of etiology. The study design, control groups, and inclusion criteria support the endpoint analyses at 1-, 3-, and 6-months post intervention based on historical data for healing, remission, and recurrence of perianal fistulas.

###### 1.3.1.2 JUSTIFICATION FOR DOSE

rhPDGF-BB is supplied in a syringe at a concentration of 0.3 mg/mL in 0.5 mL of 20 nM sodium acetate solution. Up to 4 mL, or 8 syringes (0.15-1.2 mg), of the rhPDGF-BB solution may be used to fully hydrate the collagen membrane. The volume of the rhPDGF-BB solution will be dependent on the size of the membrane needed for the fistula tract. The collagen membrane must be fully rehydrated and saturated with rhPDGF-BB for all procedures in the investigational arm of the study. The bound rhPDGF-BB will be slowly released over approximately 2-3 days into the surrounding tissue where the device is implanted. This dosing procedure and mechanism of delivery is consistent with the FDA approved devices for periodontal defects and ankle and hindfoot fusions.

###### 1.3.1.3 END OF STUDY DEFINITION

All participants in the intervention arm and participants in the control arm that heal are considered to have completed the study if he or she has finished all phases of the study including the last visit at six months shown in **section 1.1.2, Schedule of activities (SoA)**. Participants in the control arm whose treatment fails to heal the fistula at 3 months are considered to have completed the necessary endpoints for a control participant and will be offered extended open-label use of the investigational device.

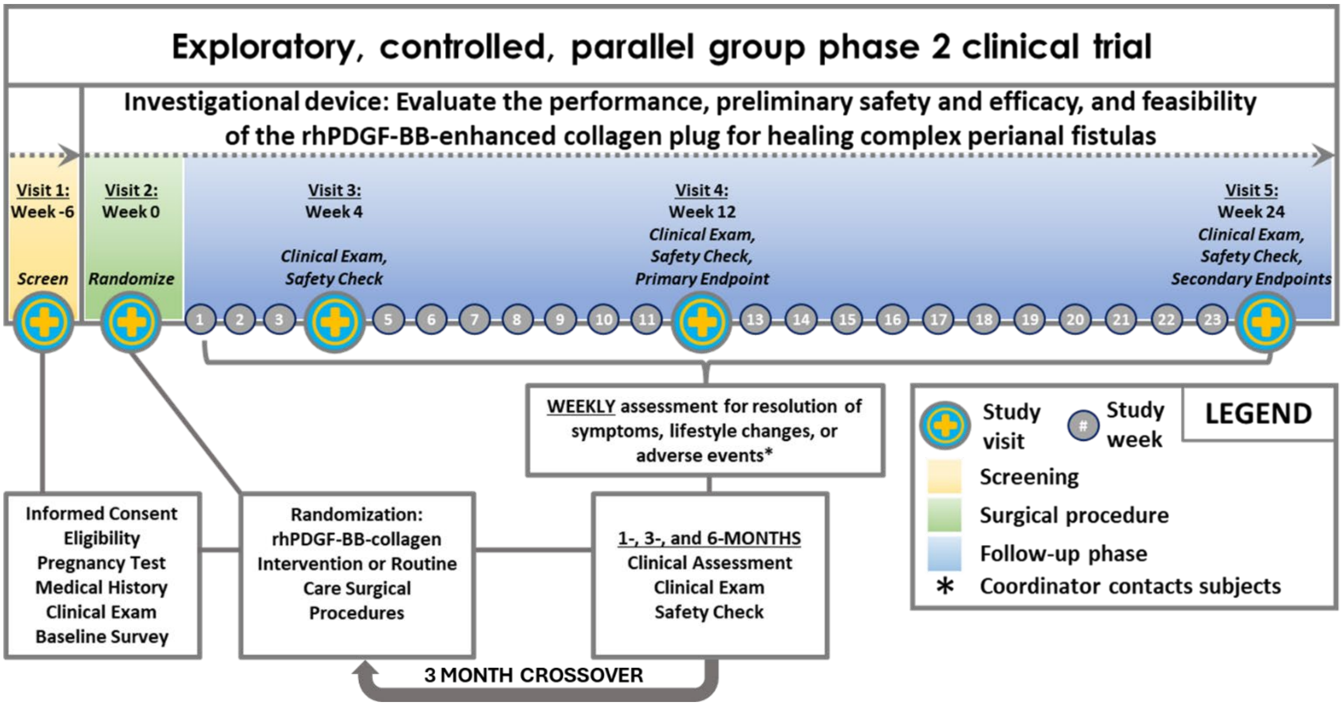

**Figure 2**. Overall Study Design

-- Confidential --
This document intended for restricted use only and may not – in full or part – be passed on, reproduced, published, or used without express permission.
All rights reserved.

#### 1.3.2 STUDY POPULATION

##### 1.3.2.1 INCLUSION CRITERIA

To be eligible to participate in this study, an individual must meet all the following criteria:

1. Diagnosis of a single tract perianal fistula not amenable to fistulotomy as determined by the supervising surgeon
2. Fistula must currently have a draining seton in place
3. Aged >21 years old
4. Willing and able to provide informed consent for study participation and compliance with study protocol
5. Stated willingness to comply with all study procedures and availability for the duration of the study

##### 1.3.2.2 EXCLUSION CRITERIA

An individual who meets any of the following criteria will be excluded from participation in this study:

1. Medical conditions that would, in the opinion of the investigator or treating provider, compromise the safety of the individual with study participation and/or the ability of the individual to follow study protocol
2. Genito-urinary fistulization, including rectovaginal (i.e., fistulas that transverse the vaginal canal)
3. Presence of an ileal anal pouch
4. Any major surgery of the gastrointestinal tract (including one or more segments of the colon or terminal ileum) within 3 months prior to randomization; presence of stoma is not exclusionary
5. Prior surgical procedure (i.e., Ligation of Intersphincteric Fistula Tract or Endorectal Advancement Flap) for the target fistula or a perianal procedure that resulted in a large soft tissue defect within 6 months prior to screening visit
6. One or more of the following fistula types or anatomic presentations: horseshoe fistulas, fistulas that do not have an opening inside the anal canal or low rectum, blind ending sinus tracts (no external opening), branching fistulas (a previously performed conversion of a branching fistula tract to a single tract is not exclusionary), >1 internal opening, moderate or severe proctitis, severe rectal mucosal fibrosis surrounding the internal opening preventing the securing of the fistula plug disk, any anatomical limitation to successfully securing the fistula plug disk.
7. Known allergic reactions to porcine collagen or yeast-derived products
8. Currently enrolled in a drug or device trial or within 30 days of last investigational drug or device administration at baseline visit where investigational treatment (drug or device) was placed in or near the fistula tract or may potentially interact with study treatment
9. Women who are pregnant, breastfeeding, or planning to become pregnant, during the trial
10. Active infection at the application site
11. The presence of malignant neoplasms at the application site
12. Prior radiation therapy at the application site

-- Confidential --
This document intended for restricted use only and may not – in full or part – be passed on, reproduced, published, or used without express permission.
All rights reserved.

###### 1.3.2.3 LIFESTYLE CONSIDERATIONS

Not applicable

###### 1.3.2.4 SCREEN FAILURES

Screen failures are defined as participants who consent to participate in the clinical trial but are not subsequently randomized or the device is unable to be implanted. A minimal set of screen failure information is required to ensure transparent reporting of screen failure participants, to meet the Consolidated Standards of Reporting Trials (CONSORT) publishing requirements and to respond to queries from regulatory authorities. Minimal information includes demography, screen failure details, eligibility criteria, and any serious adverse event (SAE).

Individuals who consent and do not meet the criteria for participation in this trial (screen failure) because of the following criteria may be rescreened. Rescreened participants should be assigned the same participant number as for the initial screening.

- Have not previously had a draining seton in place
- Have an active infection
- Surgical procedures outlined in the exclusion criteria
- Participation in an investigational study as outlined in exclusion criteria
- Pregnant or breastfeeding

###### 1.3.2.5 STRATEGIES FOR RECRUITMENT AND RETENTION

This study aims to recruit 12 participants, that are adults >21 years of age regardless of gender, race, or ethnicity. There is no information currently available regarding differential agent effects in participants defined by race or ethnicity. No vulnerable populations are expected to be enrolled in this study. The target number of patients to be screened for enrollment is 60 with the expectation of enrolling and randomizing ~25% of those screened and accounting for discontinuation, withdrawal, and loss to follow-up. This will be a single-site study at VUMC with an anticipated accrual rate of 1-2 participants per month.

Participants will be identified as potentially eligible through EMR review using a weekly EPIC export of patients scheduled to see the investigator. A list of these potential candidates will be maintained by the study coordinator in REDCap. The other surgeons in the same group as the investigator will be aware of the study may refer their patients that are potentially eligible to the investigator for the study. The Study Coordinator will review the medical records of these referrals to confirm potential eligibility and will export the phone numbers and MRNs for OK to contact patients ONLY. The patient's name, MRN, and phone number will be documented in the REDCap prescreening database. The Study Coordinator will reach out to these patients by telephone to ONLY give information about the study according to the phone script and NOT to solicit or record information about the patient to determine eligibility. If the patient does not answer, the Study Coordinator will

-- Confidential --
This document intended for restricted use only and may not – in full or part – be passed on, reproduced, published, or used without express permission.
All rights reserved.

leave a voicemail message or send a My Health At Vanderbilt message according to the scripts provided. The Study Coordinator may also try to call the patient back. This will be documented in the REDCap Prescreening database. Once reached, if the patient is interested, the Study Coordinator will schedule the initial visit in the clinic where the patient will be consented and screened. The Study Coordinator will not perform any screening activities via the phone call.

During the initial visit, the study coordinator will initiate the informed consent process. Informed consent is initiated prior to the individual's agreeing to participate in the study and continues throughout the individual's study participation. The patient consent process will be conducted using a REDCap-based electronic consent form. The consent form has been developed in REDCap, a secure, web-based, HIPAA-compliant, data collection platform with a user management system allowing project owners to grant and control varying levels of access to data collection instruments and data (e.g. read only, de-identified-only data views) for other users. Potential participants will participate in the consent process by being approached in-person at a Vanderbilt Clinic and accessing the REDCap survey via iPad or other portable electronic device. Patient signatures will be obtained using a stylus or cursor. Upon completion of the consent, patients will be provided with a copy of their version of the consent document by printing a pdf copy of the consent form in clinic. Paper consent may be used as an alternative to eConsent in case REDCap is experiencing downtime, there are issues with the electronic portable device, or the patient prefers to read and sign a paper consent form. The content between the eConsent form in REDCap and the paper consent form is identical. If paper consent is used, the patient will sign and date the form with an ink pen, and the Study Coordinator will sign and date the form as the witness. The consent form used, electronic or paper, will be documented in the patient's REDCap record. If paper consent is used, the original paper consent form will be scanned and uploaded to the patient's REDCap record in the prescreening database.

Informed Consent Forms (ICF) will be Institutional Review Board (IRB)-approved, and the participant will be asked to carefully read and review the form via an electronic device. The study coordinator will discuss benefits, risks, and alternatives of study participation with the patient. The investigator will also be available to answer any questions. Only those participants receiving the investigational device or routine care at Day 0 will be considered treated and count towards total participant population. Participants who signed consent but did not receive treatment will not be followed further. To reduce non-adherence and loss to follow-up, participants will be contacted one week prior to their scheduled follow-up visits. Communications may occur by automated phone call or e-mail message reminders, and nothing will be documented from these calls.

The following actions will be taken if a participant fails to return to the clinic for a required study visit:

1. The study coordinator will attempt to contact the participant and reschedule the missed visit prior to the next visit window and counsel the participant on the importance of maintaining the assigned visit schedule and ascertain if the participant wishes to and/or should continue in the study.
2. Before a participant is deemed lost to follow-up, the investigator or study coordinator will make every effort to regain contact with the participant (where possible, 3 telephone calls and, if necessary, a

-- Confidential --
This document intended for restricted use only and may not – in full or part – be passed on, reproduced, published, or used without express permission.
All rights reserved.

certified letter to the participant's last known mailing address). These contact attempts will be documented in the participant's medical record or study file.

3. Should the participant continue to be unreachable, he or she will be considered to have withdrawn from the study with a primary reason of lost to follow-up.

Participants will receive $200.00 for the randomized procedure and $150.00 for each follow up visit for a total of $650.00 if they complete the study.

| | |
|---|---|
| Baseline Randomized Procedure | $200 |
| Study Visits 2-4 and weekly surveys | $150/each |
| Total Reimbursement | $650 |

Participants are not allowed to accept any money for taking part in this study if they are not eligible to receive money from a U.S. person or company or the U.S. government because of U.S. national security and/or foreign policy laws. Participants can still take part in the study; however, they will not be paid if they are a resident of a country restricted by the U.S. government's comprehensive territorial sanctions or if they are listed on the U.S. Treasury Department's Office of Foreign Assets Control's Specially Designated Nationals (SDN) list of prohibited individuals. Participants do not have to say why they choose not to be paid.

##### 1.3.3 STUDY ASSESSMENTS AND PROCEDURES

###### 1.3.3.1 SAFETY ASSESSMENTS

- Clinical examination
  - At screening, during the procedure, and at the 1-month, 3-month, and 6-month follow-up visits, clinical examination of the perianal fistula will visually assess for infection, excess drainage, chronic inflammation, abnormal bleeding, or fibrosis at the external opening.
- Patient medical history
  - At screening, patients meeting inclusion criteria will be screened for exclusion criteria through their medical history. The etiology of the perianal fistula will also be discussed and documented. Participants will review any known allergies, previous surgical procedures, previous complications, and frequency of fistula occurrence or recurrence.
- Pregnancy test
  - At screening, women of childbearing potential will be required to take a pregnancy test and must agree to use two reliable contraceptive methods for the duration of their participation in the study, one of which must be a barrier method; the contraceptive methods will be documented in the screening eCRFs in REDCap
- Crohn's Concomitant Medications
  - At screening, and at the 1-month, 3-month, and 6-month follow-up visits, Crohn's medications will be documented for patients with Crohn's-related perianal fistulas, including medication name, single dose amount, route, frequency, start date, and end date.
- Participant survey
  - At screening, baseline, and weekly following the procedure till the 3-month follow-up visit, participants will complete a self-report questionnaire assessing the frequency and severity of

-- Confidential --
This document intended for restricted use only and may not – in full or part – be passed on, reproduced, published, or used without express permission.
All rights reserved.

perianal fistula-related symptoms of pain and incontinence. If symptoms are increasing in frequency and severity, an off-cycle clinical visit will be scheduled to address any adverse events.

- Assessment of adverse events
    - **Section 1.6.5**

###### 1.3.3.2 EFFICACY ASSESSMENTS

- Clinical examination
    - At screening, baseline, 1-month, 3-month, and 6-month follow-up visits, clinical examination of the perianal fistula will visually assess drainage, local inflammation, and epithelialization at the external opening to determine if the fistula is healed.
- Baseline Participant Survey
    - At consenting, participants will complete a self-report questionnaire to assess:
        - Perianal fistula-related symptoms
        - Pain scale
        - Wexner Incontinence Scale
- Follow Up Participant Survey
    - Following the baseline procedure till the 3-month follow-up visit, participants will complete a weekly self-report questionnaire to assess:
        - Perianal fistula-related symptoms
        - Pain scale
        - Incontinence (adapted from Wexner Incontinence Scale)
- Study Intervention – Baseline only
    - Seton removal, debridement of fistula tract, and implantation of the rhPDGF-BB-enhanced collagen plug
- Study Control – Baseline only
    - Seton removal and debridement of fistula tract – Routine care for Crohn's disease-related complex perianal fistulas
    - Seton removal, debridement of fistula tract, and implantation of the commercially available anal fistula plug – Routine care for idiopathic complex perianal fistulas

##### 1.3.4 STATISTICAL METHODS

###### 1.3.4.1 SAMPLE SIZE DETERMINATION

This study will aim to recruit and screen 60 eligible patients to enroll and randomize 12 participants in a 2:1 ratio. Sample size was influenced by expected confidence interval widths for the primary outcome (technical performance of the rhPDGF-BB-enhanced collagen plug for treatment of complex perianal fistulas). Assuming

-- Confidential --
This document intended for restricted use only and may not – in full or part – be passed on, reproduced, published, or used without express permission.
All rights reserved.

that all 8 participants assigned to the intervention arm will achieve successful implementation of the treatment, the lower bound of a Clopper-Pearson 95% confidence interval would be 0.63, indicating high confidence that the treatment does not have a low probability of successful implementation. Sample size was also influenced by the efficacy outcome of healing at 3 months. Based on the prior work in the literature, we assume that the routine care arm (across the expected distribution of routine care procedures) will achieve 50% healed at 3 months (2/4 healed). If 7 of the 8 intervention arm patients achieve healing, the expected lower bound of a Carlin-Louis 95% confidence interval for the differences in proportions healed would be about –0.14, indicating high confidence that the intervention does not cause substantial harm. Furthermore, if all 8 intervention arm patients achieve healing, the expected interval lower bound will be –0.01, such that there is high confidence of treatment benefit. As will be outlined in **section 1.3.4.3.2**, primary analyses will adjust for Crohn's status, which may improve statistical power/precision further.

We assumed that study dropout before 3 months will not occur, and that crossover into the intervention arm will not occur until after ascertainment of 3-month outcomes. The time-to-healing will be considered for secondary analyses and likely have slightly higher power assuming that patient-reported data is accurate and high quality, as it achieves higher granularity to assess healing than exploring healing at 3-month follow-up. Likewise, exploratory analyses including crossover patients may improve power for assessing the primary outcome. Other secondary endpoints and subgroup analyses are exploratory in nature, and we did not perform formal power analyses for these.

##### 1.3.4.2 POPULATIONS FOR ANALYSES

All randomized participants will be used in the efficacy, feasibility, and safety analyses. The sample will be drawn from VUMC, which provides care for a diverse set of fistula patients who are representative of the wider population with this ailment in the United States (US). Patients who crossover from control to intervention prior to 6 months will be excluded from analyses of 6-month outcomes and considered as additional intervention recipients in exploratory analyses.

##### 1.3.4.3 STATISTICAL ANALYSES

###### 1.3.4.3.1 GENERAL APPROACH

There will be a formal Statistical Analysis Plan (SAP). This SAP will provide detailed descriptions of all primary, secondary, and tertiary analyses, all interim and final decision thresholds, and all required documentation to ensure the reproducibility of statistical analyses. The SAP will be finalized prior to the first analysis of data, and SAP amendments (if required) will be documented before their corresponding analyses are performed. The finalized SAP will take precedence and override the statistical considerations described in this section of the master protocol (i.e., Statistical Considerations).

Categorical and binary data will be presented using counts and percentages. Continuous data will be presented using medians and interquartile ranges. All statistical tests will use an $\alpha = 0.05$ Type I error rate, using two-

-- Confidential --
This document intended for restricted use only and may not – in full or part – be passed on, reproduced, published, or used without express permission.
All rights reserved.

tailed p-values as appropriate for the specified hypotheses, but 95% confidence intervals will be given primary focus above p-values.[44] Statistical models will be adjusted for Crohn's status, and may be adjusted for additional relevant baseline covariates to improve statistical power. These covariates will be pre-specified in the formal SAP. Model assumptions will be checked using visualizations and tests as appropriate, with data transformations and alternative models considered when assumptions are dubious.

###### 1.3.4.3.2 ANALYSIS OF THE PRIMARY ENDPOINT(S)

The primary endpoint is technical performance of the treatment. This will be reported as the proportion of participants assigned to the treatment arm with successful implementation of the treatment (without SAEs). This will be reported along with a Carlin-Louis 95% confidence interval.

###### 1.3.4.3.3 ANALYSIS OF THE SECONDARY ENDPOINT(S)

The secondary efficacy endpoints will be analyzed for all randomized participants, as appropriate. Analyses will not be dependent on the findings of the primary endpoint. The first secondary endpoint considers all device-related adverse events and will be analyzed/reported in the same way as the primary endpoint.

The next secondary endpoint is fistula healing at 3 months measured as a binary variable. The probability of healing in each arm will be calculated as the proportion of randomized participants per arm that are diagnosed as completely healed by a clinical evaluation of no visible drainage from the external opening at the 3-month follow-up visit. The endpoint will be modeled as the outcome in a logistic regression model, conditioning on the treatment arm and relevant baseline covariates (including Crohn's status), as outlined in the SAP. The effect will be presented as an adjusted odds ratio with a 95% confidence interval, as well as the marginal risk ratio after performing standardization. A p-value less than 0.05 will be considered statistically significant, but interpretation will be primarily focused on the 95% confidence interval. Residuals will be inspected to inspect model fit and validity of the homoscedasticity assumption. Missing data are expected to be limited but will be handled using multiple imputation. Individuals who are lost to follow-up before 3 months will also be included via the multiple imputation procedure, considering plausible imputations conditional on their baseline and 1 month follow-up assessments, as well as weekly symptom data. The primary analysis will be intention-to-treat (ITT) based on randomized arm, as the investigational product is not available to patients who are not randomized to treatment before the 3-month outcome assessment. Supplementary Bayesian analyses will be considered to quantify probability of patient benefit.

Supplementary analysis of this secondary endpoint will consider time in weeks to achieve complete healing from baseline up to 12 weeks. This endpoint will be determined by the self-reported measure of no leakage from the weekly participant survey and will be focused on the first achievement of complete healing, as recurrence will be assessed in other outcomes. This endpoint will be analyzed using a Cox proportional hazards model, conditional on the treatment arm and Crohn's status as outlined in the SAP. The results will be presented as an adjusted hazard ratio and a marginal hazard ratio after performing standardization. Proportional hazards will not be checked with a hypothesis test, as this is typically not a recommended procedure; instead, Kaplan-

-- Confidential --
This document intended for restricted use only and may not – in full or part – be passed on, reproduced, published, or used without express permission.
All rights reserved.

Meier curves will be calculated, and plausibility of proportional hazards will be assessed visually. If proportional hazards are violated, alternative frameworks may be used, or more complex model formulations may be considered, such as models that allow for treatment effects to vary over time. Missing data are expected to be limited, but will be handled using multiple imputation, taking the longitudinal nature of surveys into account.

The next secondary endpoint is fistula recurrence at 6-months. This will be measured as the percentage of the total number of randomized participants in each arm diagnosed with recurrence of the fistula by clinical evaluation of the external opening at the 6-month follow-up visit. This is a binary endpoint which will be analyzed using the same procedures as those outlines for the binary 3-month healing endpoint.

The final secondary endpoint is the change in the severity of symptoms at 3-months (adjusted for baseline), which will be calculated using the ordinal scale for each question in the weekly participant survey. Scores will be averaged per arm and compared between arms. A composite variable will be calculated as defined in the SAP and analyzed using ordinal logistic regression conditional on treatment arm, baseline, and baseline covariates. The proportional odds assumption will be assessed by relaxing the assumption to partial proportional odds and assessing the degree of change in reported odds ratios, Results will be presented as adjusted odds ratios and missing data will be handled using multiple imputation.

Tertiary outcomes will be reported as totals or proportions, as appropriate. Analyses of proportions retained at 3-months and 6-months will be similar to those for the primary outcome.

###### 1.3.4.3.4 SAFETY ANALYSES

Safety endpoints will be analyzed using data from the weekly participant survey and from each 1- and 3-month follow-up study visit after baseline. Monitoring and reporting of safety events will be conducted continuously as described in the Data and Safety Monitoring Plan. The frequencies of adverse events and other safety endpoints will be reported along with the treatment effect on the odds of these events (i.e., the odds ratio) and associated 95% confidence intervals using the same methods outlined for primary and secondary endpoints above.

###### 1.3.4.3.5 BASELINE DESCRIPTIVE STATISTICS

Baseline binary and categorical variables will be summarized in each arm using counts and percentages. Continuous variables will be summarized in each arm using medians and interquartile ranges. Inferential statistics will not be used as the treatment arm is randomized, however, since the proposed trial size is not large, if there are strong baseline imbalances in any baseline variables, they will be added to the set of baseline adjustment variables in models if not already included. Relevant baseline outcome measures, including fistula duration and QoL will also be included in this procedure.

###### 1.3.4.3.6 PLANNED INTERIM ANALYSES

-- Confidential --
This document intended for restricted use only and may not – in full or part – be passed on, reproduced, published, or used without express permission.
All rights reserved.

This trial will not include formal interim analyses. However, safety outcomes and other data will be summarized by a blinded statistician and reviewed intermittently by the study investigators and DSMB.

###### 1.3.4.3.7 SUB-GROUP ANALYSES

Secondary endpoints will be analyzed to assess heterogeneity of treatment effect conditional on the etiology of the perianal fistula, i.e., idiopathic or Crohn's related, if the models sufficiently converge. Analysis procedures will follow those described in the primary and secondary endpoint analyses, with additional interaction terms between fistula etiology and treatment arm.

###### 1.3.4.3.8 TABULATION OF INDIVIDUAL PARTICIPANT DATA

The following demographic and baseline characteristics will be summarized for the cohort:

- Sex and gender
- Age at enrollment
- Current perianal fistula duration (months)
- Prior perianal fistulas
- Prior gastrointestinal diagnoses
- Prior abdominal surgeries

###### 1.3.4.3.9 EXPLORATORY ANALYSES

Exploratory analyses will consider the post-intervention outcomes of individuals who cross over from control to intervention. For example, an individual who is assigned control, but fails to make progress and crosses over at 4 months will have their post-crossover data included in the intervention group in exploratory analyses.

#### 1.4 RISK ANALYSIS

##### 1.4.1 RISK/BENEFIT ASSESSMENT

###### 1.4.1.1 KNOWN POTENTIAL RISKS

Regranex (becaplermin) gel (0.1 % rhPDGF-BB) is indicated for the treatment of lower extremity diabetic neuropathic ulcers that extend into the subcutaneous tissue or beyond and have adequate blood supply and is contraindicated in patients with known neoplasms at the site of application. A clinical study and post-marketing use showed that malignancies occurred distant from the site of application in REGRANEX users, but this warning was disputed and removed by the FDA in 2019 citing multiple studies that demonstrated no increased incidence of cancer or cancer mortality with Regranex gel use.[51] The effects of becaplermin exposed joints, tendons, ligaments, and bone were not established in humans, but preclinical studies in rats injected at the

-- Confidential --
This document intended for restricted use only and may not – in full or part – be passed on, reproduced, published, or used without express permission.
All rights reserved.

metatarsals demonstrated accelerated bone remodeling, fibroplasia, and mononuclear cell infiltration. Erythematous rashes occurred in 2% of patients treated with bacaplermin gel.

GEM 21S®is a synthetic grafting system of rhPDGF-BB and a synthetic calcium phosphate matrix indicated for bone and periodontal regeneration.[52] It is contraindicated in the presence of acute infections at the surgical site, malignant neoplasms at the surgical site, hypersensitivity to any components, required intraoperative soft tissue coverage is not possible, or conditions in which bone grafting is not advisable. GEM 21S® should not be used in patients that are skeletally immature.[53] The only adverse reactions reported in the literature were associated with periodontal surgical grafting procedures, not GEM 21S®. There was no clinical evidence of increased cancer incidence or mortality in GEM 21S® patents.

Augment® Bone Graft and Augment® Injectable combines rhPDGF-BB and a synthetic calcium phosphate matrix for use in bone repair and regenerative procedures of the ankle and/or hindfoot.[54] It is contraindicated in patients with hypersensitivity to the components, with active cancer, who are skeletally immature, with active infections, with metabolic disorders affecting the skeleton, who are pregnant, or when soft tissue coverage is not achievable.[55] In the safety population from a multi-center clinical study, the Augment® Bone Graft group is greater than or equal to two percentage points higher than the control group for the adverse event categories of immune system disorders, musculoskeletal and connective tissue disorders, arthralgia, pain in extremity, and nervous system disorders.[56] The key safety conclusions were that Augment® Bone Graft treated participants had overall similar rates of treatment-emergent adverse events, serious treatment-emergent adverse events, complications, and infections compared to the control group.

Although rhPDGF-BB was not genotoxic in a battery of in vitro assays and other non-clinical studies have shown no adverse findings on reproductive/development toxicity, the safety and effectiveness of rhPDGF-BB has not been established in pregnant and nursing women. Women of childbearing potential will be required to take a pregnancy test as part of the screening process for the study and must agree to use two reliable contraceptive methods for the duration of their participation in the study, one of which must be a barrier method. In the event a study participant becomes pregnant while on study, the pregnancy will be reported as information to the IRB and followed to delivery. The mother's delivery date will be tracked in the study database, and one month after the estimated due date, the study coordinator will complete the Pregnancy Outcomes Form by entering the information from the participants EMR or through a REDCap survey for the self-reporting of outcomes. The survey will inquire about any issues with the pregnancy or the presence of a congenital anomaly. If it is suspected that the investigational product results in a congenital anomaly, this will be reported as an adverse event to the IRB and the FDA.

Geistlich Nexo-Gide® Collagen Membrane is a biocompatible collagen membrane that is indicated for the management and protection of tendon injuries.[57] It is contraindicated in individuals that have a known allergy to porcine collagen or active infection at the surgical site. Geistlich Nexo-Gide® should be used with caution in patients with impaired tissue regeneration and should not be applied until bleeding is controlled.[58] The only

-- Confidential --
This document intended for restricted use only and may not – in full or part – be passed on, reproduced, published, or used without express permission.
All rights reserved.

adverse events that may be observed are those that may occur with any surgery including swelling, bleeding, hematoma, pain, and local inflammation.

###### 1.4.1.2 KNOWN POTENTIAL BENEFITS

PDGF-BB is a native protein growth factor secreted by blood platelets at sites of injury during blood clotting. *In vitro*, preclinical, and clinical studies have demonstrated PDGF-BB's potent proliferative, chemotactic, and angiogenic effects during the multifaceted and complex process of wound healing and bone repair. PDGF-BB also enhances the formation of granulation tissue during tissue repair and regeneration. rhPDGF-BB, the manufactured protein therapy, increased the incidence of complete healing of diabetic foot ulcers when used in conjunction with good ulcer care practices,[50] increased periodontal gains in patients receiving a device implantation to treat intraosseous periodontal defects,[52] and demonstrated clinical equivalency with reduced pain and improved function compared to autographs for ankle and hindfoot arthrodesis.[54] PDGF-BB's well-defined role in the recruitment of fibroblasts and pericytes, angiogenesis, and epithelialization in soft tissue are suggestive of the potential benefits of rhPDGF-BB therapy on soft tissue regrowth and stability for complete healing and remission of perianal fistulas (**Figure 1**). This investigational drug + device combo has the potential to improve the success rate for complete healing of complex perianal fistulas, reduce the recurrence rate due to reopening of the fistula tract, and avoid complications associated with surgical interventions.

###### 1.4.1.3 ASSESSMENT OF POTENTIAL RISKS AND BENEFITS

The inclusion and exclusion criteria for this study (**section 1.3.2**) were designed to minimize the risks to participants receiving rhPDGF-BB in combination with the collagen membrane for perianal fistulas in this study. Given that the intervention will be implanted at baseline and the rhPDGF-BB will be released and metabolized over the first 2-3 days, we do not anticipate any long-term risks. The plethora of human safety data that is published supports the expectation that the only risks involved with the use of rhPDGF-BB is the potential to increase acute inflammation at the site of application following intervention. There is the possibility of erythematous at the site of application due to the accumulation of blood in dilated capillaries during the initial healing phases, but these are also the primary symptoms associated with the presence of an anal fistula. The only other risks are associated with routine care procedures. The use of a commercially available collagen plug is one type of routine care procedure with a success rate of approximately 50%. The collagen plug used in the intervention arm of the study will be saturated with rhPDGF-BB to potentially enhance healing and improve the success rate. The potential benefits of this investigational intervention to improve the healing rates, time in remission, and quality of life outweigh the minimal risks in individuals with complex perianal fistulas.

#### 1.5 DESCRIPTION OF THE INTERVENTION

##### 1.5.1 STUDY INTERVENTION

###### 1.5.1.1 STUDY INTERVENTION DESCRIPTION

-- Confidential --
This document intended for restricted use only and may not – in full or part – be passed on, reproduced, published, or used without express permission.
All rights reserved.

Recombinant human platelet derived growth factor-BB (rhPDGF-BB) will be supplied in the same syringes and concentration as packaged in the GEM 21S® growth factor enhanced matrix system. rhPDGF-BB is highly purified, aseptically processed, and sterile filled into transparent glass syringes. One syringe contains a 0.5 mL solution of 0.3 mg/mL rhPDGF-BB in 20 mM sodium acetate buffer pH6+/-0.5. rhPDGF-BB should be stored at 2ºC to 8ºC, and then allowed to reach room temperature prior to administration to the patient. The rhPDGF-BB solutions supplied in GEM 21S®, Augment® Bone Graft, and Augment® Injectable systems are identical. In the case of these products, the rhPDGF-BB solution is packaged to be added to synthetic bone matrix. For this study, we will not use the bone matrix, but the rhPDGF-BB solution will instead be used to hydrate a dual surface collagen membrane.

Geistlich Nexo-Gide® is an FDA-cleared resorbable collagen membrane manufactured from porcine collagen sourced from veterinary pigs. The native collagen molecules and fiber structures are maintained, and no additional materials are added. Antigens, lipids, and non-collagenous proteins are removed through extraction and purification steps rendering the material acellular and reducing the likelihood of acute and chronic biological reactions. The collagen membranes are available in three different sizes: 20x30 mm (model 500662), 30x40 mm (model 500663), and 40x50 mm (model 500664) and can be trimmed as needed. Nexo-Gide® is indicated for tendon injuries, providing a non-constricting, protective encasement for an injured tendon. Although this collagen membrane is not currently indicated for perianal fistula, there are several other FDA-cleared anal fistula plugs manufactured from porcine that are commercially available.

Up to 4 mL of the rhPDGF-BB (0.15-1.2 mg) solution may be used to rehydrate the collagen membrane. All components of the device and sutures are biocompatible and absorbable and will only be implanted once. As the rhPDGF-BB is slowly released into the fistula tract soft tissue, cells will migrate, proliferate, and differentiate within the collagen membrane as it creates an environment that supports cell retention at the site of tissue repair.

###### 1.5.1.2 DOSING AND ADMINISTRATION

One syringe of rhPDGF-BB contains a 0.5 mL solution of 0.3 mg/mL rhPDGF-BB in 20 mM sodium acetate buffer pH6+/-0.5 for a single use dose of 0.15 mg. The collagen membranes are available in three different sizes: 20x30 mm (model 500662), 30x40 mm (model 500663), and 40x50 mm (model 500664) and can be trimmed as needed. Depending on the size membrane required to fill the fistula tract, up to 4 mL, or 8 syringes, of rhPDGF-BB will be used to rehydrate the membrane. In this study, the following process will be performed for administration of the intervention.

1. A clinical examination under anesthesia (EUA) with an anal fistula probe will be conducted to rule out active infection, anal stenosis, or active proctitis.
2. The seton will be removed, the fistula tract curetted vigorously to remove granulation tissue lining the tract, and the tract will be flushed with saline.
3. The fistula tract length and diameter and amount of the sphincter muscle involved will be assessed by the surgeon with an anal fistula probe placed from the external to the internal opening.
4. The appropriate model collagen membrane will be used based on the length of the tract.

-- Confidential --
This document intended for restricted use only and may not – in full or part – be passed on, reproduced, published, or used without express permission.
All rights reserved.

5. The collagen membrane will be cut into strips that are longer than the fistula tract to provide enough material for suturing and trimming.
6. The strips will be collected and secured by a loop to create the fistula plug
7. The plug will be saturated with the rhPDGF-BB solution in a sterile dish for at least 10 min
8. After the plug is fully saturated with rhPDGF-BB, it will be pulled through the fistula tract carefully from the internal opening to the external opening
9. The collagen plug will be sutured to the rectal mucosa at the internal opening and any portion beyond the skin level will be trimmed to sit flush with the external opening.

#### 1.6 MONITORING PROCEDURES

##### 1.6.1 KEY ROLES AND STUDY GOVERNANCE

| **Principal Investigator** | **Medical Monitor** |
|---|---|
| *Alexander T Hawkins MD MPH,*<br>*Associate Professor of Surgery* | *Evan Brittain, MD, MSc*<br>*Assoc Professor of Medicine* |
| *Vanderbilt University Medical Center* | *Vanderbilt University* |
| *1211 21st Avenue South, Suite 220* | *2525 West End, Suite 300-A* |
| *615.343.9655* | *615.322.4382* |
| ** | ** |

##### 1.6.2 SAFETY OVERSIGHT

The investigator will be responsible for data and safety monitoring. The study coordinator has responsibility for data management and further data integrity. The study coordinator will also be responsible for reporting AEs to the investigator and medical monitor as required in the Data and Safety Monitoring Plan (DSMP). An external monitor will review the informed consent, adherence to I/E criteria, AE and SAE reporting, and data quality, and any problems found will lead to a corrective action plan. Safety oversight will be under the direction of a Data and Safety Monitoring Board (DSMB) composed of 3 independent members. They will be experts/representative: clinicians, clinical trialists, and statisticians. They will not be study investigators or involved in execution of the study protocol. Furthermore, no member should have financial, proprietary, professional, or other interests that may affect impartial, independent decision-making by the DSMB. A

-- Confidential --
This document intended for restricted use only and may not – in full or part – be passed on, reproduced, published, or used without express permission.
All rights reserved.

chairperson will be selected and will be responsible for overseeing meetings, developing agendas, and being the contact person for the DSMB. The DSMB will meet at 3, 6, and 9 months to review study results and safety reports but may meet more urgently for more serious outcomes including suspected unexpected serious adverse reactions (SUSAR). The DMSB will operate under the rules of an approved charter that will be written and reviewed at the organizational meeting of the DSMB. At this time, each data element that the DSMB needs to assess will be clearly defined.

The primary charge of the DSMB is to monitor the study for participant safety. The safety and related information the DSMB will review include:

- Participant recruitment, accrual, retention, and withdrawal information
- Adverse events (AEs) and serious adverse events (SAEs)
- Any other safety-supporting data requested by the DSMB

The DSMB also reviews data related to study conduct. Data to be reviewed regarding study conduct includes: summary of protocol violations, enrollment information, noncompliance, unanticipated problems, and information concerning withdrawal of participants. The DSMB may issue recommendations regarding study conduct when concerns arise that aspects of study conduct may threaten participant safety or study integrity. The DSMB will be copied on all correspondence to the IRB or FDA and if concerns arise, the board can be assembled via conference call to discuss the pertinent issue. A summary of the DSMB findings will be provided to the IRB or FDA if needed.

##### 1.6.3 CLINICAL MONITORING

Data, safety, and enrollment monitoring is conducted to ensure that the rights and well-being of trial participants are protected, that the reported trial data are accurate, complete, and verifiable, and that the conduct of the trial is in compliance with the currently approved protocol/amendment(s), with International Conference on Harmonisation Good Clinical Practice (ICH GCP), and with applicable regulatory requirement(s).

The study team has the responsibility for the safety of the individual participants under his or her care. Participants will be monitored for adverse events during the 3 months of follow-up. For each adverse event, the site study team will classify seriousness, relatedness, expectedness, and severity. The recording and reporting of AEs depends on the classification of seriousness, relatedness, expectedness, and severity as detailed in **section 1.6.5**. The DSMB will meet at 3, 6, and 9 months to review the trial progress. A report will be generated and distributed to the DSMB prior to each meeting and will include study results, safety reports, and enrollment trackers. There will not be an interim analysis for this study.

###### 1.6.3.1 QUALITY ASSURANCE AND QUALITY CONTROL

The study team will keep accurate records to ensure that the conduct of the study is fully documented. The study team will be responsible for the regular review of the conduct of the study for verifying adherence to the

-- Confidential --
This document intended for restricted use only and may not – in full or part – be passed on, reproduced, published, or used without express permission.
All rights reserved.

protocol, and for confirming the completeness, consistency, and accuracy of all documented data and accuracy of source documentation verification. The investigator will ensure that all eCRFs and participant study files are complete for every participant. The study team will provide direct access to all trial related source data/documents and reports for the purpose inspection by local and regulatory authorities.

Following the Data and Safety Monitoring Plan (DSMP), a data monitor will review all eCRFs, and any missing data or data anomalies will be communicated to the site study team for clarification/resolution. The monitor will verify that the clinical trial is conducted, and data are generated, documented (recorded), and reported in compliance with the protocol, International Conference on Harmonisation Good Clinical Practice (ICH GCP), and applicable regulatory requirements (e.g., Good Laboratory Practices (GLP), Good Documentation Practices (GDP)). The reports of the data monitor will be submitted to the PI.

##### 1.6.3.2 DATA HANDLING AND RECORD KEEPING

###### 1.6.3.2.1 DATA COLLECTION AND MANAGEMENT RESPONSIBILITIES

Data collection is the responsibility of the clinical trial staff at the site under the supervision of the site investigator. The investigator is responsible for ensuring the accuracy, completeness, legibility, and timeliness of the data reported. All paper source documents should be completed in a neat, legible manner to ensure accurate interpretation of data. Data recorded in the electronic case report form (eCRF) derived from source documents should be consistent with the data recorded on the source documents.

Clinical data will be entered into REDCap. The data system includes password protection and internal quality checks, such as automatic range checks, to identify data that appear inconsistent, incomplete, or inaccurate. Clinical data will be entered directly from the source documents, if applicable.

###### 1.6.3.2.2 STUDY RECORDS RETENTION

Information stored in the database will be stored indefinitely.

##### 1.6.3.3 PROTOCOL DEVIATIONS

A protocol deviation is any noncompliance with the clinical trial protocol, International Conference on Harmonisation Good Clinical Practice (ICH GCP), or Manual of Procedures (MOP) requirements. The noncompliance may be either on the part of the participant, the investigator, or the study site staff. As a result of deviations, corrective actions are to be developed by the site and implemented promptly.

These practices are consistent with ICH GCP:

- 4.5 Compliance with Protocol, sections 4.5.1, 4.5.2, and 4.5.3
- 5.1 Quality Assurance and Quality Control, section 5.1.1
- 5.20 Noncompliance, sections 5.20.1, and 5.20.2.

-- Confidential --
This document intended for restricted use only and may not – in full or part – be passed on, reproduced, published, or used without express permission.
All rights reserved.

The investigator must use continuous vigilance to identify and report deviations within 5 working days of identification of the protocol deviation. All deviations must be addressed in study source documents or eCRFs. Action taken for major deviations are delineated in the Monitoring Plan and include IRB notification and may include DSMB notification..

#### 1.6.4 STUDY DISCONTINUATION/WITHDRAWAL

##### 1.6.4.1 DISCONTINUATION OF STUDY INTERVENTION

In the event a participant presents with immediate hypersensitivity reactions within 24 hours after the procedure or delayed hypersensitivity reaction within 72 hours after the procedure, the reactions will be recorded as adverse events, and the investigator will decide if the reaction is due to either the drug or the collagen plug. Based on the investigator's assessment of the reaction, the collagen plug may be removed from the fistula tract and the patient may be treated for the presenting symptoms according to usual care for hypersensitivity reactions. Symptomatic relief may be provided with short-term corticosteroids, immunomodulatory agents, or antihistamines. This study consists of a single therapeutic dose for the patient, and therefore, discontinuation of on-going study intervention does not apply.

##### 1.6.4.2 PARTICIPANT DISCONTINUATION/WITHDRAWAL FROM THE STUDY

Participants are free to withdraw from participation in the study at any time upon request. The investigator may also withdraw the participant from the clinical study at any time based on his/her medical judgment.

An investigator may discontinue or withdraw a participant from the study for the following reasons:

- Failure to heal by the 3-month follow-up visit
- Infection or abscess which requires discontinuation of the study intervention by removal of the collagen plug followed by a routine care procedure
- Unacceptable adverse event(s)
- In the judgement of the investigator, further treatment would not be in best interest of patient
- Substantial non-compliance by the patient with the requirements of the study
- The patient uses illicit drugs that may have reasonable chance of interfering with results
- Patient is lost to follow-up
- Development of intercurrent illness or situation which would affect assessments of clinical status and study endpoints to a significant degree
- Pregnancy
    - In the event a study participant becomes pregnant prior to the 3-month follow-up visit, the pregnancy will be followed to delivery and maternal and fetal outcomes will be documented.

-- Confidential --
This document intended for restricted use only and may not – in full or part – be passed on, reproduced, published, or used without express permission.
All rights reserved.

- Incarceration
- Disease progression which requires discontinuation of the study intervention
- Participant meets an exclusion criterion (either newly developed or not previously recognized) that precludes further study participation

The reason for participant discontinuation or withdrawal from the study will be recorded in the Study Withdrawal Form within the CRFs. Subjects who sign the informed consent form and are randomized but do not receive the study intervention may be replaced. Subjects who sign the informed consent form, and are randomized and receive the study intervention, and subsequently withdraw, or are withdrawn or discontinued from the study prior to the 3-month follow-up study visit, will be replaced.

###### 1.6.4.3 LOST TO FOLLOW-UP

A participant will be considered lost to follow-up if he or she fails to return for a scheduled visit and is unable to be contacted by the study site staff.

The following actions will be taken if a participant fails to return to the clinic for a required study visit:

1. The study coordinator will attempt to contact the participant and reschedule the missed visit prior to the next visit window and counsel the participant on the importance of maintaining the assigned visit schedule and ascertain if the participant wishes to and/or should continue in the study.
2. Before a participant is deemed lost to follow-up, the investigator or study coordinator will make every effort to regain contact with the participant (where possible, 3 telephone calls and, if necessary, a certified letter to the participant's last known mailing address). These contact attempts will be documented in the participant's medical record or study file.
3. Should the participant continue to be unreachable, he or she will be considered to have withdrawn from the study with a primary reason of lost to follow-up.

##### 1.6.5 ADVERSE EVENTS AND SERIOUS ADVERSE EVENTS

###### 1.6.5.1 DEFINITION OF ADVERSE EVENTS (AE)

An adverse event (AE) is defined as any untoward medical occurrence that occurs during the course of the study, whether or not the event is related to the study drug or study procedures. If a diagnosis is clinically evident (or subsequently determined), the diagnosis, rather than the individual signs and symptoms or lab abnormalities, will be recorded as the [clinical] AE.

The site study team (led by the site investigator) will evaluate AEs with respect to: (a) seriousness; (b) causality/relatedness; (c) expectedness; and (d) severity.

###### 1.6.5.1.1 DEFINITION OF SERIOUS ADVERSE EVENTS (SAE)

-- Confidential --
This document intended for restricted use only and may not – in full or part – be passed on, reproduced, published, or used without express permission.
All rights reserved.

An SAE is defined as an adverse event that, in the view of the site's study team, resulted in any of the following consequences:
[1] Death
[2] Life-threatening condition that places the participant at immediate risk of death
[3] Inpatient hospitalization or prolongation of existing hospitalization
[4] A persistent or significant incapacity or substantial disruption of the ability to conduct normal life functions or a congenital anomaly/birth defect.

Important medical events that may not result in death, be life-threatening, or require hospitalization may be considered serious when, based on medical judgment, they jeopardize participant safety or require medical or surgical intervention to prevent one of the outcomes listed in this definition.

##### 1.6.5.2 CLASSIFICATION OF AN ADVERSE EVENT

###### 1.6.5.2.1 SEVERITY OF EVENT

AEs will be graded for severity according to the *Division of AIDS (DAIDS) Table for Grading the Severity of Adult and Pediatric Adverse Events* (also known at the DAIDS AE Grading Table). For specific events that are not included in the DAIDS AE Grading Table, the generic scale listed below will be used:

| | |
|---|---|
| **Grade 1** | Events causing no or minimal interference with usual social and functional activities, and NOT raising a concern, and NOT requiring a medical intervention/ therapy. |
| **Grade 2** | Events causing greater than minimal interference with usual social and functional activities; some assistance may be needed; no or minimal medical intervention/therapy required. |
| **Grade 3** | Events causing inability to perform usual social and functional activities; some assistance usually required; medical intervention/therapy required. |
| **Grade 4** | Events causing inability to perform basic self-care functions; medical or operative intervention indicated to prevent permanent impairment, persistent disability, or death. |
| **Grade 5** | Events resulting in death. |

###### 1.6.5.2.2 RELATIONSHIP TO STUDY INTERVENTION

All AEs must have their relationship to study procedures assessed by an investigator based on the temporal relationship between the event and study procedures and his/her clinical judgment. The degree of certainty about causality will be graded using the categories below.

-- Confidential --
This document intended for restricted use only and may not – in full or part – be passed on, reproduced, published, or used without express permission.
All rights reserved.

| | |
|---|---|
| **Definitely Related** | There is clear evidence to suggest a causal relationship between a study procedure and the adverse event, and other possible contributing factors to the adverse event can be ruled out. The adverse event occurred in a plausible time relationship to a study procedure and cannot be explained by concurrent disease or other drugs or chemicals. |
| **Probably Related** | There is evidence to suggest a causal relationship between the study procedure and adverse event and the influence of other factors is unlikely. The clinical event occurs within a reasonable time after administration of the study procedure and is unlikely to be attributed to concurrent disease or other drugs or chemicals. |
| **Potentially Related** | There is some evidence to suggest a causal relationship (e.g., the event occurred within a reasonable time after administration of the trial medication). However, other factors may have contributed to the event (e.g., the participant's clinical condition, other concomitant events). Although an AE may rate only as "possibly related" soon after discovery, it can be flagged as requiring more information and later be upgraded to "probably related" or "definitely related", as appropriate. |
| **Unlikely to be related** | A clinical event whose temporal relationship to study procedures makes a causal relationship improbable (e.g., the event did not occur within a reasonable time after administration of the study intervention) and in which other drugs or chemicals or underlying disease provides plausible explanations (e.g., the participant's clinical condition, other concomitant treatments). |
| **Not Related** | The AE is completely independent of study procedures, and/or evidence exists that the event is definitely related to another etiology. There must be an alternative, definitive etiology documented by the investigator. |
| **Relatedness cannot be determined** | The relationship between study procedures and the adverse event cannot be classified with the available information. |

For the purposes of this study, an AE is considered related to study procedures if there is a "reasonable possibility" of a causal relationship between a study procedure (including the study drug) and the AE or the relationship cannot be determined; this includes events that are classified as definitely related, probably related, potentially related, or relatedness cannot be determined.

###### 1.6.5.2.3 EXPECTEDNESS

AEs that are related to study procedures or of uncertain relation to study procedures will be assessed for expectedness. An unexpected AE is defined as an AE that is not listed in the Investigator's Brochure (IB) or study protocol as an expected consequence of a study procedure (including the study drug) or is not listed at the specificity or severity listed in the IB or study protocol. Events that are described in the IB or study protocol are classified as expected **(Section 1.4)**.

###### 1.6.5.2.4 SERIOUS UNEXPECTED SUSPECTED ADVERSE REACTION (SUSAR)

-- Confidential --
This document intended for restricted use only and may not – in full or part – be passed on, reproduced, published, or used without express permission.
All rights reserved.

A SUSAR is an adverse event that meets criteria for being serious, unexpected, and related. Severity of an AE does not factor into the SUSAR criteria other than in the context of determining expectedness (an observed event that is more severe than what is described in the IB or protocol can be classified as unexpected).

##### 1.6.5.3 TIME PERIOD AND FREQUENCY FOR EVENT ASSESSMENT AND FOLLOW-UP

At scheduled study visits through the 3-month follow-up visit, all participants will have a clinical exam and data capture relevant to the outcome measures. The study team will also seek information on adverse events (AEs) by specific questioning and, as appropriate, by examination. Information on reportable AEs will be recorded in the REDCap study database as outlined in the eCRFs. The clinical course of each event will be followed until resolution, stabilization, or until it has been ultimately determined that the study treatment or participation is not the probable cause. Consistent with standard treatment for these participants, if at any point a participant experiences an emergent event, or to prevent an emergent event from occurring, or for other clinical reason, they may be treated with additional medical and/or surgical management per discretion of treating physician.

Reportable AEs/SAEs described below will be captured on the appropriate eCRF. Information to be collected includes event description, time of onset, clinician's assessment of severity, seriousness, and expectedness, relationship to study product (assessed only by those with the training and authority to make a diagnosis), and time of resolution/stabilization of the event..

Any medical condition that is present at the time that the participant is screened will be considered as baseline and not reported as an AE. However, if the study participant's condition deteriorates at any time during the study, it may be recorded as an AE.

Changes in the severity of an AE will be documented to allow an assessment of the duration of the event at each level of severity to be performed. AEs characterized as intermittent require documentation of onset and duration of each episode.

The study investigator will record AEs with start dates occurring any time after informed consent is obtained until 7 (for non-serious AEs) or 30 calendar days (for SAEs) after the 3-month follow-up visit. At each study visit through the 3-month follow-up visit, the investigator will inquire about the occurrence of AEs/SAEs since the last visit. Events will be followed for outcome information until resolution or stabilization.

###### 1.6.5.3.1 ADVERSE EVENT RECORDING AND REPORTING

AEs captured in the study database will be reviewed by the medical monitor, discussed during study team meetings, and included in the DSMB data reports. The site study team has the responsibility for the safety of the individual participants under their care. Participants will be monitored for AEs for 3 months following the procedure. For each adverse event, an investigator will classify seriousness, relatedness, expectedness, and severity.

The following categories of adverse events will be recorded as AEs in the Adverse Event case report form:

-- Confidential --
This document intended for restricted use only and may not – in full or part – be passed on, reproduced, published, or used without express permission.
All rights reserved.

- Adverse events that are classified as related to study procedures (that is, potentially, probably, or definitely related to study procedures or of uncertain relationship to study procedures), regardless of severity.
- Serious adverse events regardless of relatedness to study procedures.
- Clinical adverse events, events diagnosed as clinical conditions, with a severity grade ≥ 3, regardless of relatedness to study procedures.

All SAEs will be followed until satisfactory resolution or until the investigator deems the event to be chronic or the participant is stable. Other supporting documentation of the event may be requested by the MM, DSMB, or IRB and will be provided as soon as possible.

AEs that do not qualify for Expedited Reporting, will be recorded according to a routine reporting schedule, which includes documenting the event on the Adverse Event eCRF within 14 calendar days of site awareness of the event. The DSMB will review all recorded AEs during scheduled meetings. The SPM will distribute the written summary of the DSMB's review including the review of AEs to the IRB. If the DSMB determines the overall rate of AEs is higher in the intervention group than the control group, the SPM will notify the IRB and the FDA within 14 calendar days of this determination.

##### 1.6.5.3.2 EXPEDITED REPORTING

The following events will be reported using an expedited reporting schedule:

- AEs that are related and serious are reported to the IRB and DSMB
- AEs that are related, unexpected, and grade ≥ 3 are reported to the IRB and DSMB
- SUSARs (AEs that are related, serious, and unexpected) are reported to the IRB, DSMB, and FDA

###### 1.6.5.3.2.1 AES THAT ARE RELATED AND EITHER SERIOUS OR UNEXPECTED AND GRADE > 3 (BUT NOT BOTH)

AEs that are either serious or unexpected and grade ≥ 3 will be recorded and an assessment of whether there is a reasonable possibility that study procedures caused the event will be conducted and if still determined to be potentially, probably, or definitely related or relatedness cannot be determined, the event will be reported to the IRB and DSMB as soon as possible but not later than 7 calendar days after site awareness of the event.

All serious adverse events (SAEs) will be followed until satisfactory resolution or until the investigator deems the event to be chronic or the participant is stable. Other supporting documentation of the event may be requested by the medical monitor, DSMB, or IRB and should be provided as soon as possible.

###### 1.6.5.3.2.2 SERIOUS UNEXPECTED SUSPECTED ADVERSE REACTION (SUSAR) REPORTING

The Food and Drug Administration (FDA) must be notified of any unexpected fatal or life-threatening suspected adverse reaction as soon as possible, but in no case later than 7 calendar days after the investigator's initial receipt of the information.  In addition, the FDA must be notified in an Investigational New Drug (IND)

-- Confidential --
This document intended for restricted use only and may not – in full or part – be passed on, reproduced, published, or used without express permission.
All rights reserved.

safety report of potential serious risks, from clinical trials or any other source, as soon as possible, but in no case later than 15 calendar days after it is determined that the information qualifies for reporting.

###### 1.6.5.3.3 REPORTING EVENTS TO PARTICIPANTS

Not applicable

###### 1.6.5.3.4 EVENTS OF SPECIAL INTEREST

Not applicable

###### 1.6.5.3.5 REPORTING OF PREGNANCY

In the event a study participant becomes pregnant while on study, the pregnancy will be reported as information to the IRB and followed to delivery. The mother's delivery date will be tracked in the study database, and one month after the estimated due date, the study coordinator will complete the Pregnancy Outcomes Form by entering the information from the participants EMR or through a REDCap survey for the self-reporting of outcomes. The survey will inquire about any issues with the pregnancy or the presence of a congenital anomaly. If it is suspected that the investigational product results in a congenital anomaly, this will be reported as an adverse event to the IRB and the FDA.

##### 1.6.5.4 UNANTICIPATED PROBLEMS

###### 1.6.5.4.1 DEFINITION OF UNANTICIPATED PROBLEMS (UP)

The Office for Human Research Protections (OHRP) considers unanticipated problems involving risks to participants or others to include, in general, any incident, experience, or outcome that meets all of the following criteria:

- Unexpected in terms of nature, severity, or frequency given (a) the research procedures that are described in the protocol-related documents, such as the Institutional Review Board (IRB)-approved research protocol and informed consent document; and (b) the characteristics of the participant population being studied;
- Related or possibly related to participation in the research ("possibly related" means there is a reasonable possibility that the incident, experience, or outcome may have been caused by the procedures involved in the research); and
- Suggests that the research places participants or others at a greater risk of harm (including physical, psychological, economic, or social harm) than was previously known or recognized.

-- Confidential --
This document intended for restricted use only and may not – in full or part – be passed on, reproduced, published, or used without express permission.
All rights reserved.

###### 1.6.5.4.2 UNANTICIPATED PROBLEM REPORTING

Unanticipated problems (UPs) will be reported to the reviewing Institutional Review Board (IRB). The UP report will include the following information:

- Protocol identifying information: protocol title and number, PI's name, and the IRB project number;
- A detailed description of the event, incident, experience, or outcome;
- An explanation of the basis for determining that the event, incident, experience, or outcome represents an UP;
- A description of any changes to the protocol or other corrective actions that have been taken or are proposed in response to the UP.

To satisfy the requirement for prompt reporting, UPs will be reported using the following timeline:

- UPs that are serious adverse events (SAEs) will be reported to the IRB within 7 calendar days of the investigator becoming aware of the event.
- Any other UP will be reported to the IRB within 15 calendar days of the investigator becoming aware of the problem

###### 1.6.5.4.3 REPORTING UNANTICIPATED PROBLEMS TO PARTICIPANTS

Not applicable

#### 1.7 MANUFACTURING INFORMATION

##### 1.7.1 ACQUISITION AND ACCOUNTABILITY

The rhPDGF-BB and Geistlich Nexo-Gide® will be donated by Lynch Regenerative Medicine and Geistlich Pharma North America, Inc. rhPDGF-BB will be labeled as [CAUTION-Investigational New Drug. Limited by Federal (or United States) Law to Investigational Use] on the outer packaging and stored in a 4°C refrigerator that is controlled and monitored by Dr. Yan Ru Su's Core Lab for Translational & Clinical Research at VUMC. The investigator or a delegate will be responsible for acquiring the rhPDGF-BB from the laboratory on the day of a procedure. The collagen membranes will be labeled as [CAUTION-Investigational Device. Limited by Federal (or United States) Law to Investigational Use] and stored at room temperature in a secure location designated for investigational devices. The investigator or a delegate will be responsible for acquiring the collagen membranes from the secure location on the day of a procedure. All unused or expired products will be returned to Lynch Regenerative Medicine for proper disposal.

##### 1.7.2 FORMULATION, APPEARANCE, PACKAGING, AND LABELING

-- Confidential --
This document intended for restricted use only and may not – in full or part – be passed on, reproduced, published, or used without express permission.
All rights reserved.

rhPDGF-BB will be supplied as packaged in the GEM 21S® growth factor enhanced matrix system, in a clear glass syringe with 0.5 mL of 0.3 mg/mL rhPDGF-BB in 20 mM sodium acetate buffer. The outer box packaging includes the product name, contents and concentration, expiration date, and manufacturer. A sticker will be placed on the outer packaging indicating [CAUTION-Investigational New Drug. Limited by Federal (or United States) Law to Investigational Use].

One Geistlich Nexo-Gide® collagen membrane and one aluminum template are separately packed in sterile inner blisters and packaged in a single outer blister. The outer box label includes the product name, contents, size of membrane, and manufacturer. A sticker will be placed on the outer box indicating [CAUTION-Investigational Device. Limited by Federal (or United States) Law to Investigational Use].

##### 1.7.3 PRODUCT STORAGE AND STABILITY

rhPDGF-BB:

- Must be refrigerated at 2° to 8° C (36° to 46° F)
- Protected from light prior to use
- Do not remove from outer covering prior to use

Geistlich Nexo-Gide® collagen membrane:

- Store at controlled room temperature 59°–77 °F (15°–25 °C) and in a dry place.
- Keep away from sunlight.
- The device should be handled using sterile gloves and sterile instruments.

##### 1.7.4 PREPARATION

1. The rhPDGF-BB will be removed from 4°C storage and allowed to sit at room temperature while the fistula is being prepared and examined.
2. The appropriate model collagen membrane will be selected based on the length of the tract.
3. The collagen membrane will be cut into strips that are longer than the fistula tract to provide enough material for suturing and trimming.
4. The strips will be saturated with the rhPDGF-BB solution in a sterile dish for a minimum of 10 min before use.

###### 1.7.4.1 MEASURES TO MINIMIZE BIAS: RANDOMIZATION AND BLINDING

To prevent bias in allocation of participants to arms, participant eligibility should be confirmed prior to releasing the randomization allocation. Randomization will occur at the baseline visit at a ratio of 2:1 intervention to control. Randomization will be performed through the REDCap randomization module, which allows for loading a randomization table and creates a "Randomize" button for each participant. Randomization

-- Confidential --
This document intended for restricted use only and may not – in full or part – be passed on, reproduced, published, or used without express permission.
All rights reserved.

will be stratified to achieve balance in treatment allocation across Crohn's vs. idiopathic fistula types. Because the total sample size is 12, randomization will be blocked using blocks of size 3 (2:1 intervention to control) within each stratum (Crohn's vs idiopathic). Blinding of the patients, surgeons other than the investigator performing the procedure, and study team members assessing outcomes to study arm allocation will be employed to reduce bias in conducting study activities and evaluations. Given that the control group will be receiving a routine care procedure, the investigator who will be performing all procedures will be unblinded to the treatment assignment for each participant. Prior to the scheduled procedure, the investigator will use the randomization module in REDCap to determine the assignment of the participant. On the day of the procedure, the investigator or a delegate will retrieve the rhPDGF-BB from the laboratory and the collagen membrane and take it to the operating room for participants randomized to the investigational treatment group. To maintain blinding while assessing outcomes, clinical exams during follow-up study visits will be performed by another surgeon not involved in the initial screening visit or the procedural encounter. In the case of a serious adverse event or medical emergency in which knowledge of the treatment is necessary for the participant's well-being or medical care, the unblinded investigator will be alerted to ensure appropriate medical management.

###### 1.7.4.2 STUDY INTERVENTION COMPLIANCE

This study consists of a single therapeutic dose via implanted device for the patient, and therefore, study intervention compliance does not apply.

###### 1.7.4.3 CONCOMITANT THERAPY

Concomitant systemic treatments for Crohn's disease will be guided by the patient's clinicians according to standard of care. If the patient has a Crohn's exacerbation and the treating gastroenterologist wants to increase immunomodulation therapy, we as a study team will not attempt to prevent that. However, systemic treatment for the CD-related fistula will not be initiated or changed during the study. The study team will document all Crohn's-related treatments in the study database.

###### 1.7.4.3.1 RESCUE MEDICINE

Hypersensitivity to rhPDGF-BB has not been previously observed in humans. Skin irritation, vasodilation, central nervous system depression, and accelerated bone remodeling was observed after intravenous dosing in mice. Porcine collagen is most similar to human collagen, so it has almost no immune restriction. Multiple studies assessing hypersensitivity in humans demonstrated that porcine collagen did not cause foreign body reactions or significant erythematous reactions, and the porcine collagen membranes were biocompatible. In the event a participant presents with immediate hypersensitivity reactions within 24 hours after the procedure or delayed hypersensitivity reaction within 72 hours after the procedure, the reactions will be recorded as adverse events, and the investigator will decide if the reaction is due to either the drug or the collagen plug. Based on the investigator's assessment of the reaction, the collagen plug may be removed from the fistula tract and the patient may be treated for the presenting symptoms according to usual care for hypersensitivity reactions.

-- Confidential --
This document intended for restricted use only and may not – in full or part – be passed on, reproduced, published, or used without express permission.
All rights reserved.

Symptomatic relief may be provided with short-term corticosteroids, immunomodulatory agents, or antihistamines.

### 1.8 SUPPORTING DOCUMENTS AND OPERATIONAL CONSIDERATIONS

#### 1.8.1 REGULATORY AND ETHICAL CONSIDERATIONS

##### 1.8.1.1 INFORMED CONSENT PROCESS

###### 1.8.1.1.1 CONSENT/ASSENT AND OTHER INFORMATIONAL DOCUMENTS PROVIDED TO PARTICIPANTS

Consent forms describing in detail the study intervention, study procedures, and risks are given to the participant and written documentation of informed consent is required prior to administering study intervention.

###### 1.8.1.1.2 CONSENT PROCEDURES AND DOCUMENTATION

Participants will be identified as potentially eligible through EMR review using a weekly EPIC export of patients scheduled to see the investigator. A list of these potential candidates will be maintained by the study coordinator in REDCap. The other surgeons in the same group as the investigator will be aware of the study and may refer their patients that are potentially eligible to the investigator for the study. The Study Coordinator will review the medical records of these referrals to confirm potential eligibility and will export the phone numbers and MRNs for OK to contact patients ONLY. The patient's name, MRN, and phone number will be documented in the REDCap prescreening database. The Study Coordinator will reach out to these patients by telephone to ONLY give information about the study according to the phone script and NOT to solicit or record information about the patient to determine eligibility. If the patient does not answer, the Study Coordinator will leave a voicemail message or send a My Health At Vanderbilt message according to the scripts provided. The Study Coordinator may also try to call the patient back. This will be documented in the REDCap Prescreening database. Once reached, if the patient is interested, the Study Coordinator will schedule the initial visit in the clinic where the patient will be consented and screened. The Study Coordinator will not perform any screening activities via the phone call.

Informed consent is initiated prior to the individual's agreeing to participate in the study and continues throughout the individual's study participation. The patient consent process will be conducted using a REDCap-based electronic consent form. The consent form has been developed in REDCap, a secure, web-based, HIPAA-compliant, data collection platform with a user management system allowing project owners to grant and control varying levels of access to data collection instruments and data (e.g. read only, de-identified-only data views) for other users. Potential participants will participate in the consent process by being approached in-person at a Vanderbilt Clinic and accessing the REDCap survey via iPad or other portable electronic device. Patient signatures will be obtained using a stylus or cursor. Upon completion of the consent, patients will be

-- Confidential --
This document intended for restricted use only and may not – in full or part – be passed on, reproduced, published, or used without express permission.
All rights reserved.

provided with a copy of their version of the consent document by printing a pdf copy of the consent form in clinic. Paper consent may be used as an alternative to eConsent in case REDCap is experiencing downtime, there are issues with the electronic portable device, or the patient prefers to read and sign a paper consent form. The content between the eConsent form in REDCap and the paper consent form is identical. If paper consent is used, the patient will sign and date the form with an ink pen, and the Study Coordinator will sign and date the form as the witness. The consent form used, electronic or paper, will be documented in the patient's REDCap record. If paper consent is used, the original paper consent form will be scanned and uploaded to the patient's REDCap record in the prescreening database.

Informed Consent Forms (ICF) will be Institutional Review Board (IRB)-approved, and the participant will be asked to carefully read and review the form via an electronic device or the paper form. The Study Coordinator will explain the research study, discuss benefits, risks, and alternatives of study participation, and answer any questions that may arise. The Primary Investigator will also be available to answer any questions. A verbal explanation will be provided in terms suited to the participant's comprehension of the purposes, procedures, and potential risks of the study and of their rights as research participants. Participants will have the opportunity to carefully review the form and ask questions prior to signing. The participants will have the opportunity to discuss the study with their family and think about it prior to agreeing to participate. Participants will be informed that participation is voluntary and that they may withdraw from the study at any time, without prejudice. The informed consent form will be signed before the participant undergoes any study-specific procedures. A copy of the informed consent document will be given to the participants for their records. The rights and welfare of the participants will be protected by emphasizing to them that the quality of their medical care will not be adversely affected if they decline to participate in this study. The Study Coordinator will complete the consent attestation after consent is obtained.

###### 1.8.1.2 STUDY DISCONTINUATION AND CLOSURE

This study may be temporarily suspended or prematurely terminated if there is sufficient reasonable cause determined by the DSMB. Written notification, documenting the reason for study suspension or termination, will be provided by the suspending or terminating party to the study participants, investigator, and regulatory authorities. If the study is prematurely terminated or suspended, the Principal Investigator (PI) will promptly inform study participants and the Institutional Review Board (IRB) and will provide the reason(s) for the termination or suspension. Study participants will be contacted, as applicable, and be informed of changes to study visit schedule.

Circumstances that may warrant termination or suspension include, but are not limited to:

- Determination of unexpected, significant, or unacceptable risk to participants
- Demonstration of efficacy that would warrant stopping
- Insufficient compliance to protocol requirements
- Data that are not sufficiently complete and/or evaluable
- Determination that the primary endpoint has been met
- Determination of futility

-- Confidential --
This document intended for restricted use only and may not – in full or part – be passed on, reproduced, published, or used without express permission.
All rights reserved.

Study may resume once concerns about safety, protocol compliance, and data quality are addressed, and satisfy the IRB and/or Food and Drug Administration (FDA).

###### 1.8.1.3 CONFIDENTIALITY AND PRIVACY

Participant confidentiality and privacy is strictly held in trust by the study team, and all research activities will be conducted in as private a setting as possible. This confidentiality is extended to cover the participant's clinical information and the study protocol, documentation, data, and all other information generated. No information concerning the study or data will be released to any unauthorized third party. All faculty and staff working on the study receive and provide documentation of training in Good Clinical Practice (GCP) as part of their onboarding and continuing education.

Upon signing consent, each participant will be assigned a unique study participant identification number (PTID). Clinical data from this study, including participant birthdate and demographics, will be associated with the PTID and maintained on a Health Insurance Portability and Accountability Act (HIPAA) compliant Research Electronic Data Capture (REDCap) database accessible to the study team only. REDCap is a secure, web-based application designed to support data capture for research studies, providing 1) an intuitive interface for validated data entry; 2) audit trails for tracking data manipulation and export procedures; 3) automated export procedures for seamless data downloads to common statistical packages; and 4) procedures for importing data from external sources. Hard copy clinical data forms, for example source or protocol-specific Case Report Forms (CRFs), associated with the PTID will be scanned directly into REDCap. Afterwards, the paper copy will be placed in designated secure bins for shredding documents with protected health information on Vanderbilt's campus. The hard copy may be saved as needed in the secure cabinet in the locked research office. A file linking the participant personal identifiable information (PII) (i.e., their name and contact information) to their PTID will be maintained in the REDCap database. Study personnel will only send documents containing participant PII via fax or encrypted email in accordance with HIPAA regulations (e.g., to send/receive safety lab data and medical records request to/from primary care provider).

Project team members listed as Key Study Personnel with existing electronic health record (EHR) system access rights may also be granted use of REDCap Clinical Data Interoperability Services (CDIS) tools. These tools are designed to enable transfer of relevant study-related data from the Vanderbilt Research Derivative and/or directly from the EHR into REDCap.

Representatives of the Institutional Review Board (IRB) or regulatory agencies may inspect all documents and records required to be maintained by the investigator, including but not limited to, medical records (office, clinic, or hospital) and pharmacy records for the participants in this study. The clinical study site will permit access to such records.

The study participant’s contact information will be securely stored within the REDCap database for internal use during the study. At the end of the study, all records will continue to be housed in REDCap indefinitely.

###### 1.8.1.4 FUTURE USE OF STORED SPECIMENS AND DATA

-- Confidential --
This document intended for restricted use only and may not – in full or part – be passed on, reproduced, published, or used without express permission.
All rights reserved.

Not applicable

###### 1.8.1.5 PUBLICATION AND DATA SHARING POLICY

This study will be conducted in accordance with the following publication and data sharing policies and regulations:

This study will comply with the NIH Data Sharing Policy and Policy on the Dissemination of NIH-Funded Clinical Trial Information and the Clinical Trials Registration and Results Information Submission rule. As such, this trial will be registered at ClinicalTrials.gov, and results information from this trial will be submitted to ClinicalTrials.gov. In addition, every attempt will be made to publish results in peer-reviewed journals.

###### 1.8.1.6 CONFLICT OF INTEREST POLICY

The independence of this study from any actual or perceived influence, such as by the pharmaceutical industry, is critical. Therefore, any actual conflict of interest of persons who have a role in the design, conduct, analysis, publication, or any aspect of this trial will be disclosed and managed. Furthermore, persons who have a perceived conflict of interest will be required to have such conflicts managed in a way that is appropriate to their participation in the design and conduct of this trial. The study leadership has established policies and procedures for all study group members to disclose all conflicts of interest and will establish a mechanism for the management of all reported dualities of interest.

##### 1.8.2 ADDITIONAL CONSIDERATIONS

Not applicable

##### 1.8.3 PROTOCOL AMENDMENT HISTORY

| Version | Date | Description of Change | Brief Rationale |
|---|---|---|---|
| V.2 | 19 March 2024 | Updated protocol for IND Potential hold comments | FDA sent potential hold comments and requests for information |
| V.3 | 26 March 2024 | Updated for IRB application | IRB application reviewed by experts in VUMC |
| V.4 | 29 April 2024 | Updated for IRB pre-review | Regulatory compliance analyst reviews the IRB submission prior to the IRB committee review |

-- Confidential --
This document intended for restricted use only and may not – in full or part – be passed on, reproduced, published, or used without express permission.
All rights reserved.

| | | | |
|---|---|---|---|
| V.5 | 30 May 2024 | Updated for IRB Committee Recommendations | The IRB committee decided to defer our application until we respond to their recommendations |
| V.6 | September 2024 | IRB Amendments: SoA updates, eConsent, AE recording and reporting, | Schedule of activities updated to reflect the surveys and safety monitoring through the 3-month follow-up visit, adding eConsent as an alternative to paper, updated AE recording and reporting procedures |
| V.6.1 | 13 December 2024 | IRB Amendment: Add paper consent as a backup method and add messaging through MHAV to contact potential participants | To add paper consent as a backup in case there are issues with the portable electronic device or the patient prefers paper and to add My Health at Vanderbilt as an approved contact method for reaching out to potential participants who have been referred to Dr. Hawkins' study by his colleagues. In past experience, patients are more responsive to messages from MHAV than voicemail. |

-- Confidential --
This document intended for restricted use only and may not – in full or part – be passed on, reproduced, published, or used without express permission.
All rights reserved.

-- Confidential --
This document intended for restricted use only and may not – in full or part – be passed on, reproduced, published, or used without express permission.
All rights reserved.
