## Supplementary material for "Study protocol and statistical analysis plan for a randomized controlled trial evaluating the safety and feasibility of the recombinant human platelet-derived growth factor B (rhPDGF-BB)-enhanced collagen plug for complex perianal fistula healing": S3 Appendix

### VUMC Institutional Review Board
### Informed Consent Document for Research

| | |
|---|---|
| **Study Title:** | **A randomized controlled trial evaluating the safety and feasibility of the recombinant human platelet-derived growth factor B (rhPDGF-BB)-enhanced collagen plug for complex perianal fistula healing** |
| **Version** | **6.1** |
| **Date:** | **13 January 2025** |
| **PI:** | **Alexander Hawkins, MD** |

Name of participant: ___________________________________________ Age: ___________

**The following is given to you to tell you about this research study. Please read this form with care and ask any questions you may have about this study. Your questions will be answered. Also, you will be given a copy of this consent form.**

**Key Information:**
The first section of this document contains some key points that the research team thought you would find important. The study is described in more detail after this section. If you do not understand something, please ask someone.

**<u>Key information about this study:</u>**
The purpose of this study is to determine if a healing factor called recombinant human platelet-derived growth factor-BB (rhPDGF-BB) delivered on a collagen plug can improve the healing of complex perianal fistulas. rhPDGF-BB is similar to a protein that is naturally made in your body at sites of injury and is FDA-approved for healing other types of skin and tissue wounds. rhPDGF-BB has been safe for humans in more than 100 clinical trials and for over 20 years of real-world use. For this study, rhPDGF-BB will be applied to the fistula channel, which is a new location in the body where we are studying how it may benefit healing. If you decide to take part in this study, you will be randomly assigned to one of two groups: the investigational rhPDGF-BB-enhanced collagen plug or the routine care procedure. This study will assign 2 participants to the investigational group for every 1 participant that is assigned to the routine care group. This is called a 2:1 randomization and means that double the number of participants will receive the healing factor versus routine care.

This box is for
IRB USE ONLY
Do not edit or delete

| | |
|---|---|
| **Study Title:** | **A randomized controlled trial evaluating the safety and feasibility of the recombinant human platelet-derived growth factor B (rhPDGF-BB)-enhanced collagen plug for complex perianal fistula healing** |
| **Version** | **6.1** |
| **Date:** | **13 January 2025** |
| **PI:** | **Alexander Hawkins, MD** |

**Detailed Information:**
The rest of this document includes detailed information about this study (in addition to the information listed above).

**Detailed information about this study:**
You are being asked to take part in this research study because you have been diagnosed with a perianal fistula that is idiopathic (unknown cause) or due to Crohn's disease, and your surgeon does not believe a fistulotomy is the best option for you. When a fistulotomy is not an option, surgeons will consider other routine care treatments. One of these is a collagen plug, which is a cone-shaped plug made of connective tissue that is placed in the fistula to promote tissue repair. In some situations, it may be determined that allowing the fistula to heal naturally is the most suitable course of action. The purpose of this study is to investigate whether a specific type of collagen plug enhanced with the healing factor, rhPDGF-BB, is a better option than the existing procedures.

You do not have to be in this research study. You may choose not to be in this study and get other treatments without changing your healthcare, services, or other rights. You can stop being in this study at any time. If we learn something new that may affect the risks or benefits of this study, you will be told so that you can decide whether or not you still want to be in this study. Your medical record will contain a note saying you are in a research study and may contain some research information about you. Anyone you authorize to receive your medical record will also get this information.

**Side effects and risks that you can expect if you take part in this study:**
**Side effects of rhPDGF-BB include:**
**Common:** Swelling, redness, and tenderness of the skin at the application site.
**Rare but serious side effects:** Allergic reactions to this product are possible. Patients with known allergies to collagen derived from pigs or yeast-derived products should not use this

This box is for
IRB USE ONLY
Do not edit or delete

product. **Risks that are not known:** Because this treatment is investigational, meaning non-FDA approved, there may be risks that we do not know about at this time.

**Women of childbearing potential:** The safety and effectiveness of the healing factor, rhPDGF-BB, has not been established in pregnant and nursing women. If you are pregnant or currently breast feeding, you may not participate in this study. Please know that if you are pregnant, if you become pregnant, or if you are breast-feeding during this study, you or your child may be exposed to an unknown risk. To confirm to the extent medically possible that you are not pregnant, you must have a pregnancy test done before beginning this research study. You must also agree to use two reliable contraceptive methods for the duration of participation in the study, one of which must be a barrier method.

Contraception Methods:

- Barrier method of contraception: condoms with or without a spermicidal agent, diaphragm, or cervical cap with spermicide
- IUD
- Hormone-based contraceptive

You must accept the risk that pregnancy could still result despite the responsible use of reliable method of birth control. You must notify the investigator as soon as possible of any failure of proper use of your birth control method, or if you become pregnant, either of which may result in your being withdrawn from the study. In the event you become pregnant while on study, the pregnancy will be followed to delivery and outcomes will be documented via your electronic medical record or self-reported survey.

**Good effects that might result from this study:**

The benefits to science and humankind that might result from this study: If this treatment is effective, it will provide a better option for treating perianal fistulas.

This box is for
IRB USE ONLY
Do not edit or delete

**Procedures to be followed:**
The purpose of this study is to see whether at a collagen plug enriched with the healing factor, rhPDGF-BB, can improve the healing of complex perianal fistulas.

You are being asked to participate in a randomized study. This means that you will be randomly assigned to receive either the collagen plug with the healing factor or receive routine care treatment, an anal fistula plug or natural healing. After the procedure, we will continue to check in on you through in-person visits at 1 month, 3 months, and 6 months. At each follow up visit, we will discuss your progress, perform a routine care clinical examination, and take a clinical photograph of the perianal area. We will discuss any adverse events at the 1-month and 3-month follow-up visits. As another way of keeping track of your healing progress, we will ask you to fill out weekly online health questionnaires at baseline and weekly starting at 1 week after the procedure. These weekly questionnaires will continue until week 12 (about 3 months) after the procedure.

If you are randomized to the routine care treatment group and your fistula is not healed at 3 months, you will be offered the option to crossover to an open-label use group and receive the collagen plug with the healing factor. After the intervention procedure, you will restart at baseline, and we will continue to check in on you through in-person visits at 1 month, 3 months, and 6 months and ask you to fill out weekly online health questionnaires starting at 1 week after the procedure. These weekly questionnaires will continue until week 12 (about 3 months) after the procedure.

There may be alternative procedures or courses of treatment that may be advantageous to your clinical care. At the discretion of your supervising surgeon, if one of these alternatives is thought to be clearly better for your specific situation, then you may be asked to withdraw from the study.

**Payments for your time spent taking part in this study or expenses:**

This box is for
IRB USE ONLY
Do not edit or delete

You will be reimbursed for taking part in this study. You will receive $200.00 for the randomized procedure and $150.00 for each follow up visit for a total of $650.00 if you complete the study.

| | |
|---|---|
| Study visit 1 – Randomized Procedure | $200 |
| Study visits 2-4 – Follow up exams | $150/each |
| Total Reimbursement | $650 |

You are not allowed to accept any money for taking part in this study if you are not eligible to receive money from a U.S. person or company or the U.S. government because of U.S. national security and/or foreign policy laws. You can still take part in the study; however, you will not be paid if you are a resident of a country restricted by the U.S. government‘s comprehensive territorial sanctions or if you are listed on the U.S. Treasury Department’s Office of Foreign Assets Control’s Specially Designated Nationals (SDN) list of prohibited individuals. You do not have to say why you choose not to be paid.

**Costs to you if you take part in this study:**
If you agree to take part in this research study, you and/or your insurance will not have to pay for the tests and treatments that are being done only for research. However, you are still responsible for paying for the usual care you would normally receive for the treatment of your illness. This includes treatments and tests you would need even if you were not in this study. These costs will be billed to you and/or your insurance.

You have the right to ask what it may cost you to take part in this study. If you would like assistance, financial counseling is available through the Vanderbilt Financial Assistance Program. The study staff can help you contact this program. You have the right to contact your insurance company to discuss the costs of your routine care (non-research) further before choosing to be in the study. You may choose not to be in this study if your insurance does not pay for your routine care (non-research) costs and your doctor will discuss other treatment plans with you.

This box is for
IRB USE ONLY
Do not edit or delete

**Payment in case you are injured because of this research study:**
If it is determined by Vanderbilt and the Investigator that an injury occurred, then you and/or your insurance may be billed for the cost of medical care provided at Vanderbilt to treat the injury. You will be responsible for any copayments or deductibles associated with the treatment of that injury.

There are no plans for Vanderbilt or the Sponsor to pay for the costs of any additional care. There are no plans for Vanderbilt or the Sponsor to give you money for the injury.

**Who to call for any questions or in case you are injured:**
If you should have any questions about this research study or if you feel you have been hurt by being a part of this study, please feel free to contact Alex Hawkins, MD at 615-322-2063.

For additional information about giving consent or your rights as a person in this study, to discuss problems, concerns, and questions, or to offer input, please feel free to call the VUMC Institutional Review Board Office at (615) 322-2918 or toll free at (866) 224-8273.

**Reasons why the study doctor may take you out of this study:**
Your surgeon may withdraw you from the study at any time based on his/her medical judgment. You may be withdrawn from the study if you no longer meet the inclusion criteria, you meet an exclusion criterion, or if your surgeon believes that a different procedure would be better for you. In the case of any adverse reaction to the study drug, your study doctor may decide to take you out of the study early.

**What will happen if you decide to stop being in this study?**

This box is for
IRB USE ONLY
Do not edit or delete

If you decide to stop being part of the study, you should tell your study doctor. Deciding to not be part of the study will not change your regular medical care in any way.

**Clinical Trials Registry:**
A description of this clinical trial will be available on www.clinicaltrials.gov, as required by U.S. Law. This website will not include information that can identify you.

**Confidentiality:**
Your records associated with this study will be kept in an electronic database called REDCap that is designed to safely store sensitive health information. Only the principal investigator and key study personnel will have access to this information. Before analyzing the results, all information that could potentially be used to link a research record with an individual's identity will be removed. These measures are designed to protect your health information, but as with any study there is a risk that confidentiality will be lost.

This confidentiality does not protect information that we have to report by law, such as child abuse or some infectious diseases. It does not prevent us from disclosing your information if we learn of possible harm to yourself or others, or if you need medical help.

Disclosures that you consent to in this document are not protected. This includes putting research data in the medical record or sharing research data for this study or future research. Disclosures that you make yourself are also not protected.

**Privacy:**
Only approved key study personnel will have access to your health information associated with the study. Approved key study personnel may access your electronic medical record periodically only to collect information that is relevant for initial inclusion or exclusion in the study as well as in follow-up after the procedure. Because this study involves a drug combination device, the Food and Drug Administration may inspect records associated with the study.

*This box is for*
IRB USE ONLY
*Do not edit or delete*

**Study Results:**
The aggregate results of the study will be shared with the financial sponsor, Lynch Regenerative Medicine, the scientific community through a scientific manuscript, and the public through a summary on www.clinicaltrials.gov. These results will not contain any identifiable personal information. The financial sponsor may also receive a de-identified dataset (which will not contain any identifiable personal information).

**Authorization to Use/Disclose Protected Health Information:**
**What information is being collected, used, or shared?**
To do this research, we will need to collect, use, and share your private health information. By signing this document, you agree that your health care providers (including both Vanderbilt University Medical Center and others) may release your private health information to us, and that we may use any and all of your information that the study team believes it needs to conduct the study. Your private information may include things learned from the procedures described in this consent form, as well as information from your medical record (which may include information such as HIV status, drug, alcohol or STD treatment, genetic test results, or mental health treatment).

**Who will see, use, or share the information?**
The people who may request, receive, or use your private health information include the researchers and their staff. Additionally, we may share your information with other people at Vanderbilt, for example if needed for your clinical care or study oversight. By signing this form, you give permission to the research team to share your deidentified information with others outside of Vanderbilt University Medical Center. This may include the financial sponsor of the study and its agents or contractors, outside providers, study safety monitors, government agencies, other sites in the study, data managers and other agents and contractors used by the study team. We try to make sure that everyone who sees your information keeps it confidential, but we cannot guarantee that your information will not be shared with others. If your information is disclosed by your health care providers or the research team to others, federal and state confidentiality laws may no longer protect it.

This box is for
IRB USE ONLY
Do not edit or delete

| | |
|---|---|
| **Study Title:** | **A randomized controlled trial evaluating the safety and feasibility of the recombinant human platelet-derived growth factor B (rhPDGF-BB)-enhanced collagen plug for complex perianal fistula healing** |
| **Version** | **6.1** |
| **Date:** | **13 January 2025** |
| **PI:** | **Alexander Hawkins, MD** |

**Do you have to sign this Authorization?**
You do not have to sign this Authorization, but if you do not, you may not join the study.

**How long will your information be used or shared?**
Your Authorization for the collection, use, and sharing of your deidentified information does not expire. Additionally, you agree that your deidentified information may be used for similar or related future research studies.

**What if you change your mind?**
You may change your mind and cancel this Authorization at any time. If you cancel, you must contact the Principal Investigator in writing to let them know by using the contact information provided in this consent form. Your cancellation will not affect information already collected in the study, or information that has already been shared with others before you cancelled your authorization.

**If you decide not to take part in this research study, it will not affect your treatment, payment or enrollment in any health plans or affect your ability to get benefits. You will get a copy of this form after it is signed.**

This box is for
IRB USE ONLY
Do not edit or delete

#### STATEMENT BY PERSON AGREEING TO BE IN THIS STUDY

**I have read this consent form, and the research study has been explained to me verbally. All my questions have been answered, and I freely and voluntarily choose to take part in this study.**

____________________ ____________________________________________

Date/Time Signature of patient/volunteer

Consent obtained by:

____________________ ____________________________________________

Date/Time Signature

____________________________________________

Printed Name and Title

This box is for
IRB USE ONLY
Do not edit or delete
